# Detection and characterisation of copy number variants from exome sequencing in the DDD study

**DOI:** 10.1101/2023.08.23.23294463

**Authors:** Petr Danecek, Eugene J. Gardner, Tomas W. Fitzgerald, Giuseppe Gallone, Joanna Kaplanis, Ruth Y. Eberhardt, Caroline F. Wright, Helen V. Firth, Matthew E. Hurles

**Affiliations:** Wellcome Sanger Institute, Wellcome Genome Campus, Hinxton, UK; Department of Clinical and Biomedical Sciences, University of Exeter Medical School, Royal Devon and Exeter Hospital, Exeter, United Kingdom; Department of Clinical Genetics, Cambridge University Hospitals NHS Foundation Trust, Cambridge, UK

## Abstract

**Purpose:** Structural variants such as multi-exon deletions and duplications are an important cause of disease, but are often overlooked in standard exome/genome sequencing analysis. We aimed to evaluate the detection of copy number variants (CNVs) from exome sequencing (ES) in comparison to genome-wide low-resolution and exon-resolution chromosomal microarrays (CMA), and to characterise the properties of *de novo* CNVs in a large clinical cohort.

**Methods:** We performed CNV detection using ES of 13,462 parent-offspring trios in the Deciphering Developmental Disorders (DDD) study, and compared them to CNVs detected from exon-resolution array comparative genomic hybridization (aCGH) in 5,197 probands from the DDD study.

**Results:** Integrating calls from multiple ES-based CNV algorithms using random forest machine learning generated a higher quality dataset than using individual algorithms. Both ES- and aCGH-based approaches had the same sensitivity of 89% and detected the same number of unique pathogenic CNVs not called by the other approach. Of DDD probands pre-screened with low resolution CMA, 2.6% had a pathogenic CNV detected by higher resolution assays. *De novo* CNVs were strongly enriched in known DD-associated genes and exhibited no bias in parental age or sex.

**Conclusion:** ES-based CNV calling has higher sensitivity than low-resolution CMAs currently in clinical use, and comparable sensitivity to exon-resolution CMA. With sufficient investment in bioinformatic analysis, exome-based CNV detection could replace low-resolution CMA for detecting pathogenic CNVs.

## Introduction

Copy Number Variants (CNVs) are differences in the number of copies of genomic segments and constitute a class of genetic variation of both medical and evolutionary importance. It has been estimated that 3-14% of patients with rare developmental disorders (DDs) harbour a pathogenic CNV^1,2-3^. This diagnostic yield varies depending on the assay used for detection, the patient cohort and clinical history of the patients^4-5^. Most pathogenic CNVs have a dominant mode of inheritance and many arise *de novo* in DD patients. While population surveys have shown that the mutational mechanisms generating CNVs generate a continuous spectrum of CNV sizes, with smaller CNVs outnumbering larger CNVs^6^, among pathogenic CNVs large CNVs predominate. This contrast is partly due to larger CNVs being more likely to be pathogenic, and partly due to the low sensitivity of detecting smaller CNVs in clinical testing.

In most clinical genetics services, the primary assay used to identify CNVs in DD patients is chromosomal microarray (CMA) analysis. A range of different CMA assays are in widespread use, which vary in the size of CNVs that can be detected due to differences in the number, placement and performance of the individual oligonucleotide probes on a given array. Many of these CMA assays use a genome-wide backbone of probes (often 60,000-180,000 probes) with the aim of detecting CNVs that are hundreds of kilobases in size or larger – here termed ‘low-resolution’ CMA. Some CMA assays augment this genomic backbone with probes targeting exons of known dosage-sensitive genes in order to increase sensitivity to detect small pathogenic CNVs; however, CMA assays that target all protein-coding exons are not in widespread use.

As more clinical genetics services adopt next-generation sequencing for diagnosing DD, it would be both advantageous and cost-effective to determine all classes of pathogenic genetic variation from this single assay. Both exome and genome sequencing interrogate all protein-coding exons, and therefore, in principle, can detect the complete size range of CNVs impacting these exons. However, compared to small insertions/deletions (InDels) and single nucleotide variants (SNVs), CNVs and structural variation in general remain more difficult to detect with high accuracy^6,7^. A number of approaches have been developed^8^, but no comprehensive assessment comparing the specificity and sensitivity of exon-resolution CMA versus exome sequencing (ES)-based approaches has been performed in very large patient cohorts.

Here we compare CNV detection from ES and genome-wide exon-resolution CMA in the DDD study cohort of 13,462 probands with severe DD recruited from the United Kingdom and Republic of Ireland, of which 9,859 probands were sequenced as parent-offspring trios^9^. Additionally, 5,197 probands have exon-resolution CMA data available from a custom array comprising 1.11M probes targeting 97% of protein coding exons of GENCODE transcripts (v2010-07-22)^10^ with multiple probes per protein-coding exon and an additional 0.67M genomic backbone probes in non-coding regions to give an overall median probe spacing of 2kb (Additional File 1). Most (73%) but not all of the 9,859 DD probands were pre-screened by low-resolution CMA prior to recruitment to the DDD study, allowing us to assess the added diagnostic yield of exon-level resolution in CNV detection. We also present a systematic characterisation of 598 *de novo* CNVs detected in the trios.

## Materials and methods

### Sample and data collection

Children with severe, undiagnosed developmental disorders were recruited for the DDD study by 24 centers across the UK and Ireland as previously described^11^. The study performed genome-wide aCGH on blood-derived DNA and ES of saliva- or blood-derived DNA. Among the 13,462 probands with ES data, 9,859 were sequenced as parent-offsping trios and of these 5,197 had both proband aCGH and parent-offspring trio ES data available. Prior to enrolment into DDD 7,182 (73%) of the 9,859 participants were pre-screened using clinical-grade CMA (typically a low-resolution array) to exclude patients with large pathogenic CNVs or aneuploidy.

### CNV calling from aCGH data

The custom CGH microarray (AMADID array design IDs: 031220/031221) used for CNV discovery was designed to have good power to detect single exon CNVs by using 5 probes per exon, as well as having a dense backbone of ~320k intronic and intergenic probes with a median probe spacing of 2kb. It is composed of two 1 million probe Agilent arrays and has been designed to target genes and ultra-conserved elements throughout the human genome. The aCGH CNV calls were obtained using a custom algorithm, CNsolidate (Supplement S1).

### CNV calling from ES data

The ES data were generated using one of two custom bait designs based on the Agilent SureSelect v3 and v5 exomes, supplemented with additional probes targeting ~6,000 high value non-coding regions comprising 12.9% of the total targeted sequence^12^. The sequencing was performed as described previously^1^. The paired-end 75 bp sequencing reads were aligned onto the GRCh37d5 reference genome with bwa^13^. Four programs were used to generate the initial raw callset: CANOES^14^, CLAMMS^15^, CoNVex^16^, and XHMM^17^; the calls were then integrated using a random forest machine learning approach (Supplements S2-S4), their breakpoints resolved (Supplements S2 and S21), their inheritance status determined from overlapping parental calls, and their parent of origin ascertained from informative SNVs (Supplement S18).

### Gene Enrichment Testing

To test enrichment in genes associated with developmental disorders, we compiled a subset of 800 genes in which *de novo* mutations are robustly associated with disease (i.e. monoallelic and X-linked) from the Developmental Disorders Genotype-To-Phenotype Database (DDG2P^18^, https://www.ebi.ac.uk/gene2phenotype/downloads/DDG2P.csv.gz version 2022-10-17) restricted to categories “definitive” or “strong”. In further text we refer to these genes in which de novo mutation is sufficient to cause DD simply as the “DN-DD genes”. For details see Supplement S6.

### Clinical Evaluation of Potentially Pathogenic CNVs

CNVs were selected for clinical review by a DDD clinical reporting pipeline prior to this work ^11^ and independently of the random forest classification described above. All CNVs discovered by CNsolidate and CoNVex were filtered based on annotations provided by the callers, CNV size, overlap with common and recurrent calls, and overlap with DDG2P genes (Supplement S5). A set of 276 clinically validated pathogenic *de novo* CNVs from samples with both exon-resolution CMA and ES data available was used as a truth set to assess sensitivity of the methods.

### Code Availability

All code is freely available (Supplement S3).

## Results

### Modelling anticipated sensitivity of different CNV detection assays

Previous studies have suggested that CNV detection assays typically require at least two or three probes (or baits for exome sequencing) within the CNV for accurate detection. Therefore, to establish approximate baseline expectations of the sensitivity of different CNV detection assays to detect CNVs of different sizes (drawn from a population-based CNV size distribution established by the 1000 genomes project, see Supplement S7) we performed simulations of the sensitivity of four different CNV detection assays: low-resolution CMA with either 60K (CytoSure Constitutional v3) or 180K (Agilent CGH ISCA v2) probes, the exon-resolution 2M CMA designed specifically for the DDD study, and exome sequencing (Supplement S7). The custom exon-resolution CMA is expected to have the highest sensitivity because it targets most protein-coding exons with at least five probes. These simulations suggested that if we assumed that two probes/baits are sufficient for reliable CNV detection then exon-resolution CMA would be expected to have 98% sensitivity for single exon CNVs, whereas exome sequencing (which uses a single bait for most exons) would have 31% sensitivity, and the 180K and 60K CMA platforms would have 39% and 17% sensitivity, respectively. However, for CNVs which affect more than 3 exons, both exon-resolution CMA and ES-based CNV ascertainment would be expected to have 99% sensitivity, which exceeds by a wide margin the anticipated sensitivity of low resolution CMA (39% and 69%; Supplement S8a). We note that the 180k CMA design that we modelled includes probes targeting exons of known dosage-sensitive genes^19^ and would be expected to detect 89% of simulated CNVs impacting DD-associated genes, compared to 60% by the 60k array and 99.9% by the 2M CMA and ES (Supplement S8b).

### CNV ascertainment in probands with severe developmental disorders

Four ES-based CNV calling algorithms (CANOES^14^, CLAMMS^15^, CoNVex^16^, and XHMM^17^) were run on 32,523 DDD proband and parental samples (Methods; Supplement S2). To ascertain CNVs from the 5,197 probands with exon-resolution CMA data, we used the CNsolidate algorithm (Supplement S1). We observed differences in the number of calls between the four ES CNV callers despite the same underlying data (Supplement S9), ranging from an arithmetic mean of 15 calls per sample from XHMM to 67 calls per sample from CoNVex. The initial union ES-based CNV callset consisted of 9.6M calls with unique breakpoints within a sample, but included possible duplicate calls with discrepant breakpoints produced by different programs. After merging potentially redundant calls (Supplement S2), 7.3M non-redundant CNV calls remained (Supplement S10a).

To integrate the four ES-based CNV callers and generate a combined callset of improved accuracy, we annotated each ES-based CNV call with a range of quality-related metrics and trained a random forest classifier on 1,332 common CNVs (Supplement S2). We trained separate random forest models for deletions and copy number gains (in further text referred to as “duplications”), the most informative variables differed between the two models (Supplementary Figure S23). In total, 54,607 calls (0.75%) passed the random forest filtering threshold (Supplement S11) of which 718 (1.3%) were not observed in parents of the trios (i.e. were putative *de novo* CNVs). We further excluded 120 (17%) of these putative *de novo* CNVs because 15 (2%) were in regions of the genome that are known to rearrange in blood cell lineages and 82 (11%) were also observed at implausibly high frequencies (N>24) in 17,208 unrelated, unaffected individuals (parents of other trios) and were thus unlikely to be pathogenic (the threshold of N=24 was set based on prevalence of known pathogenic recurrent CNV syndromes in the same set of individuals^20^). A further 23 CNVs (3%) were observed in healthy control samples in public databases of structural variation (N>24 in DGV^21^ or AF>0.01 in 1000 Genomes Project^22^ or GnomAD-SV^6^) and are thus also unlikely to be pathogenic. The final ES-based callset used in subsequent analyses thus consisted of 598 *de novo* rare CNVs in 13,462 probands (Supplement S10b).

Comparison of the individual and combined ES-based CNV callsets to a truth set of 276 clinically validated pathogenic *de novo* CNVs largely discovered from the DDD exon-resolution CMA showed that the accuracy of all individual ES-CNV callers was considerably lower than that of the combined random forest callset (Figure 1). For example, if controlling false positive calls equally by restricting each callset to a maximum of 0.15 putative *de novo* CNVs called per sample^4-5^, the sensitivity to *de novo* CNVs of individual ES callers would be between 10-68% for duplications and 26-65% for deletions. In comparison, the predicted sensitivity of the integrated random forest callset at this stringent filtering threshold was 84% for both duplications and deletions. Note that variant quality scores reported by some of the programs are capped at the higher end, thus truncating the lower range of expected number of variants per sample in Figure 1. For example, even though CLAMMS was the most sensitive caller with respect to the 276 clinically validated pathogenic *de novo* CNVs (Additional File 2), the top bin of its quality score distribution contained more calls with an identical quality score than the other callers, which means that quality score filtering alone cannot create a higher-specificity callset closer to the expected number of *de novo* CNVs (Supplement S24).

**Figure 1.**
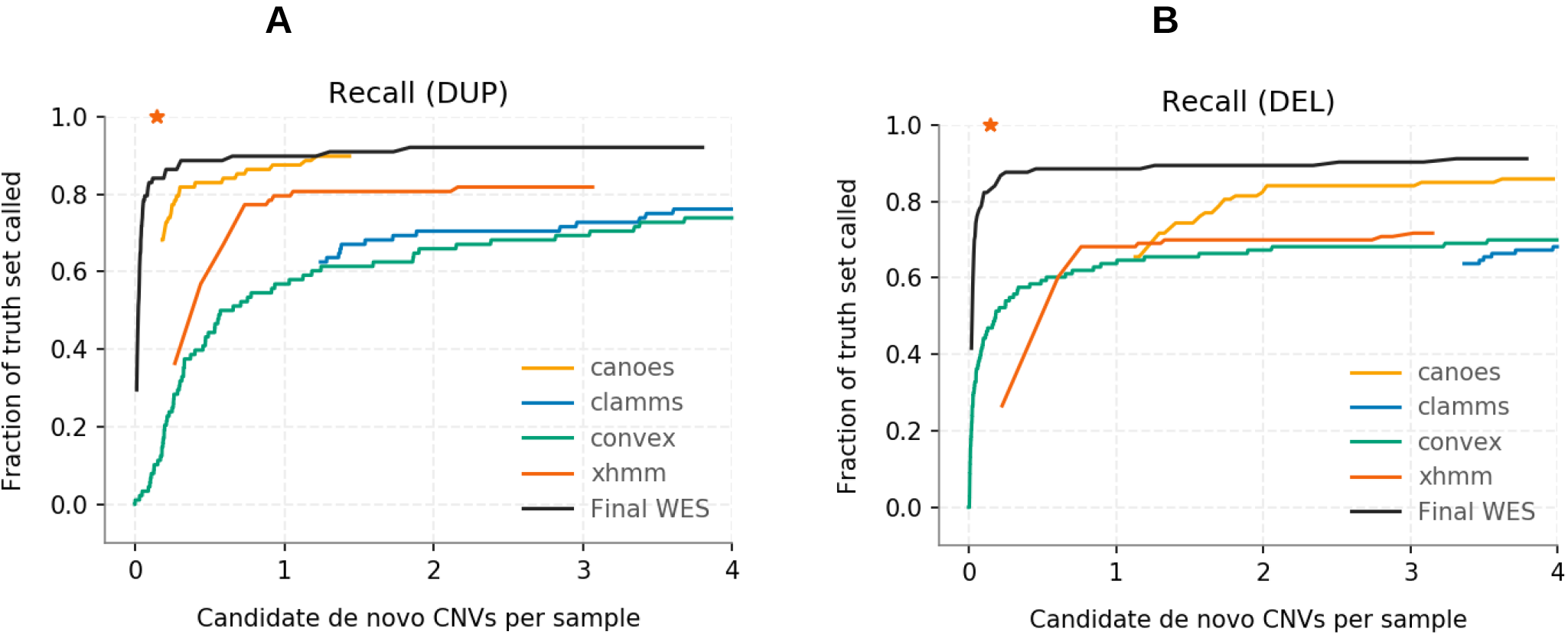
Pseudo-ROC curves showing sensitivity of individual WES callers and our final callset versus the number of de novo calls per sample under varying quality cutoffs separately for duplications (**A**) and deletions (**B**). The truth set consists of 276 clinically validated pathogenic CNVs discovered mainly from the exon-resolution CMA. The red asterisk in each plot denotes 0.15 putative de novo CNVs called per sample.

We next assessed the overall sensitivity against 276 clinically validated pathogenic CNVs identified in these individuals (largely from the exon-resolution CMA). The random forest integrated callset identified 246 (89%) of these CNVs. For large CNVs (i.e. >10 exome baits) ES-based ascertainment was at least as sensitive as exon-resolution CMA; filtered ES-based calls achieved 98% sensitivity while the sensitivity of exon-resolution CMA calls was only 92% at the applied thresholds. In general, most of the pathogenic CNVs missed by ES (18/30, 60%) intersected either one (9/30) or zero (9/30) exome baits, and are thus inaccessible to discovery (Supplement S12). While most pathogenic CNVs missed by exon-resolution CMA or ES-based calling had small (<10) numbers of probes/baits, a few larger CNVs (4/30 of those missed by ES-based calling) were missed due to being fragmented into smaller number of calls, none of which met the required quality thresholds. In comparison, the exon-resolution CMA also identified 246 (89%) of the 276 clinically validated pathogenic CNVs.

### Characteristics of *de novo* CNVs

Among the 598 high quality *de novo* CNVs we detected, *de novo* deletions were more frequent than duplications (64% vs 36%), consistent with other large scale studies^6,23^. Sixty-five percent (N=391) of *de novo* CNVs impacted the coding sequence of multiple genes while thirty-two percent (N=194) impacted the coding sequence of a single gene (Supplement S13), and three percent (N=13) did not impact coding sequence. Among single-gene CNVs, only 10% overlapped all coding exons of the gene; most were partial-gene CNVs. Approximately half of *de novo* CNVs intersected DD-associated genes in which *de novo* mutations can be sufficient to cause disease (DN-DD genes) (Figure 2a). This proportion is much higher than the proportion of exome baits that target exons of DN-DD genes (6.6%), and is much higher than the proportion of inherited CNVs that encompass DN-DD genes, either in population studies, or in the DDD families. A permutation test (Supplement S6), assuming a uniform genome-wide CNV mutation rate, confirmed that DDD probands have significantly more *de novo* deletions and duplications which impact DN-DD genes (2.0x for duplications and 3.0x for deletions; p < 1e-10) than expected by chance. The same test also showed that *de novo* deletions and duplications are significantly enriched in a broader set of constrained genes (1.2x and 1.5x; p = 2.6e-3 and p < 1e-10) with high probability of intolerance to heterozygous loss of function variants^24^ (pLI > 0.9) (Supplement S14), but not in biallelic DD-associated genes or in genes that are not DD-associated (Figure 2b).

**Figure 2.**
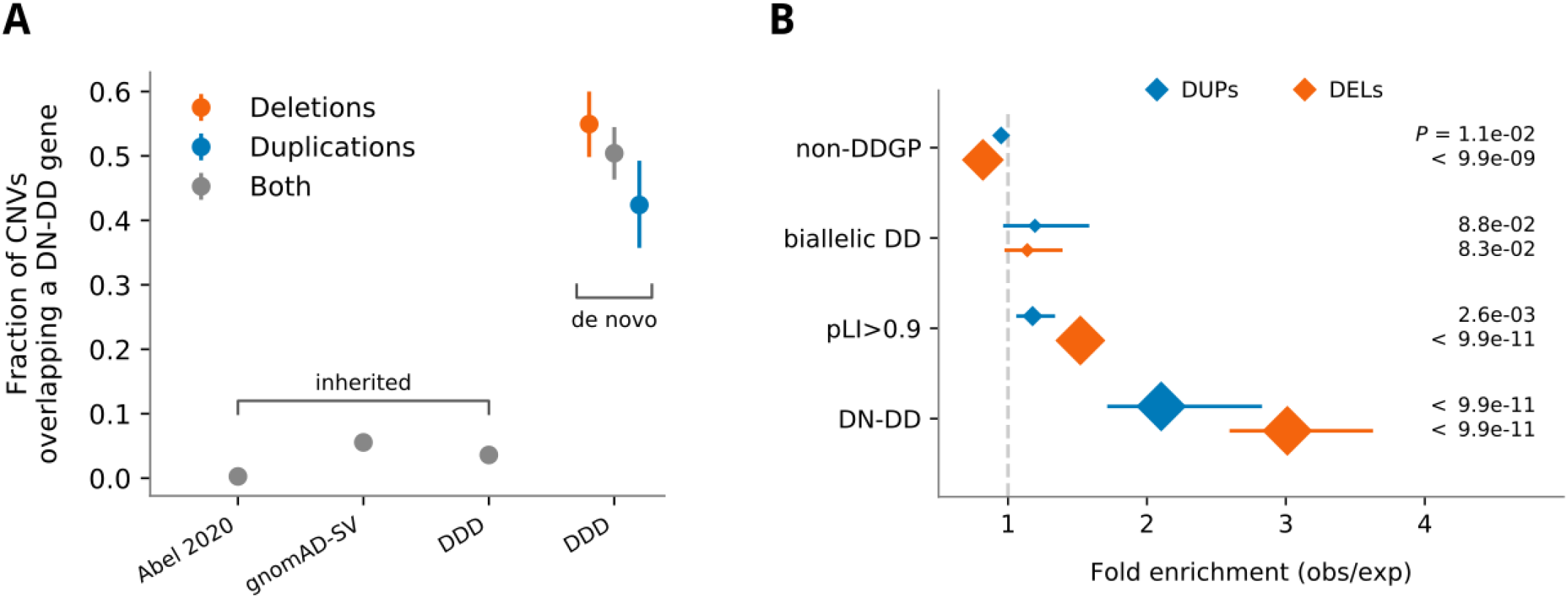
**A**. Fraction of de novo CNVs overlapping a DN-DD gene compared to inherited CNVs in the DDD and two other large scale studies. **B**. Enrichment of de novo CNVs in genes in which a de novo mutation is sufficient to cause a disease (DN-DD), in constrained genes with high pLI score (pLI>0.9), in recessive biallelic genes (biallelic DD), and in genes not previously associated with developmental disease (non-DDGP). The intervals show 90% of the simulated distribution (Supplement S14) and the size of the diamonds indicates the significance of the results with P values shown on the right.

The enrichment of de novo duplications in DD-associated genes could be driven by triplosensitivity or by gene-disrupting duplications. Among duplications impacting a single gene, we observed that a higher proportion of partial gene duplications (28%, 13/46) than entire gene duplications (9%, 1/11) impacts DN-DD genes (Supplement S15). This is in stark contrast to deletions impacting a single gene, where we observed a lower proportion of partial gene deletions (51%, 66/129) than entire gene deletions impacting DN-DD genes (88%, 7/8). This suggests that an appreciable proportion of pathogenic *de novo* duplications are likely to be gene disrupting rather than operating via triplosensitivity.

To investigate genes specifically associated with DDs caused by CNVs, we first looked at single-gene CNVs. Twenty-eight genes were impacted recurrently by the 195 *de novo* single-gene CNVs, of which 20 were known DN-DD genes (Figure 3a). Of the remaining eight genes, two (PUM1, SPG7) are associated with other neurological monogenic disorders in OMIM (www.omim/org) and two (*TANC2, PTPRT*) already have partial but inconclusive evidence of association with neurodevelopmental disorders^25,26^.

**Figure 3.**
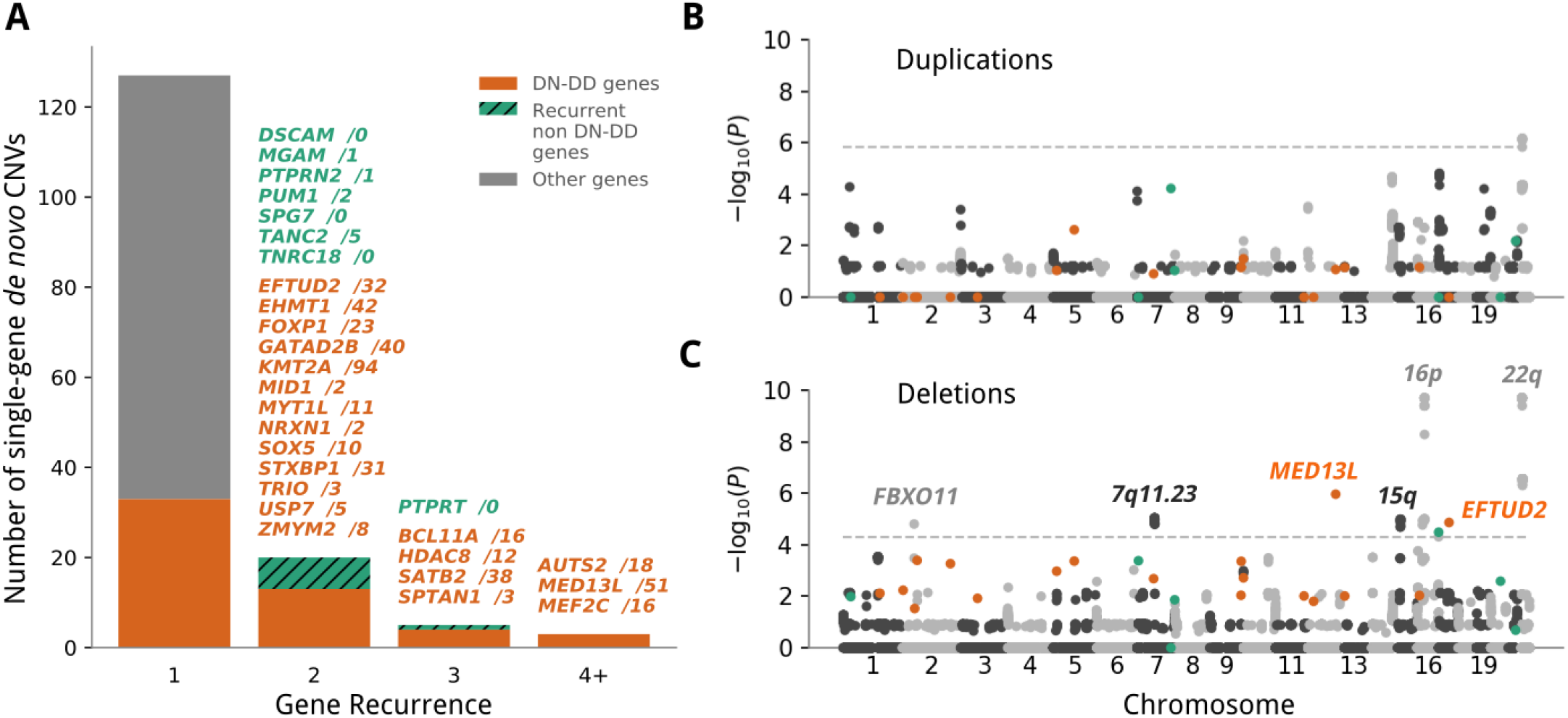
**A**. Gene recurrence in the 158 de novo CNVs which overlapped the coding sequence of a **single** gene. The gene names are followed by the number of samples with a protein truncating variant in DD patients from a recent analysis of 31,058 trios^29^; most of the genes that were affected multiple times across the DDD patients were either known or novel candidate DD genes in the study. **B-C**. Gene recurrence in de novo CNVs overlapping coding sequence of any number of genes separately for duplications (**B**) and deletions (**C**). Highlighted in grey/black text are known neurodevelopmental loci which harbour statistically significant genes according to our test (Supplement S15). P values were determined by a permutation test which consisted of 5e9 iterations each under an assumption of a uniform CNV rate. The dashed line marks a false discovery rate of 0.01 as calculated by the Benjamini-Hochberg procedure.

We then tested for gene-specific enrichment of CNVs across the complete set of 598 *de novo* CNVs, including those impacting multiple genes, using a genome-wide permutation test assuming a uniform CNV rate (Methods). In total, 168 genes passed genome-wide significance threshold (Benjamini-Hochberg corrected p < 0.01; Figure 3b-c); 10 genes (Supplement S16a) were known DN-DD genes^18^. Of the remaining 158 genes which passed genome-wide significance, all but one are located within or flanking (±1 Mbp) known DD-associated recurrent pathogenic CNVs (Supplement S16b)^27^. The remaining significant gene, *SPG7*, has previously been associated with autosomal recessive spastic paraplegia^28^. To assess if *SPG7* is a potential DN-DD gene candidate we performed in-depth clinical review of all three patients with deletions intersecting *SPG7*. Two of three *de novo SPG7* deletions also overlap either the coding sequence or promoter of the flanking well-characterized DN-DD gene *ANKRD11* and both patients presented with phenotypes consistent with *ANKRD11* loss, while the third patient also has a *de novo* protein-truncating SNV in the gene *KMT2A* consistent with their symptoms. As such, we consider *SPG7* to be a likely passenger alongside the pathogenic partial deletions of *ANKRD11*.

### Association of *de novo* CNVs with Parental Age and Sex

We next sought to determine if there was any parental bias in the origin of *de novo* CNVs (Supplement S17-18). A strong paternal bias has been observed for other classes of variation (e.g. ~80% for SNVs^30^ and ~75% for InDels^31^) but the evidence for CNVs has been mixed; previous studies have observed strong, weak, or absent paternal bias^32,33,34^ but also a strong maternal bias at specific loci or for aneuploidies^35,36^. We were able to determine parental origin for 360 (64%) of *de novo* autosomal CNVs, 189 (53%) of which had paternal origin, a non-significant bias (binomial test p = 0.40; Figure 4, Supplement S19). There were no obvious parental biases when stratifying these CNVs into deletions and duplications or larger and smaller events (Figure 4a).

**Figure 4.**
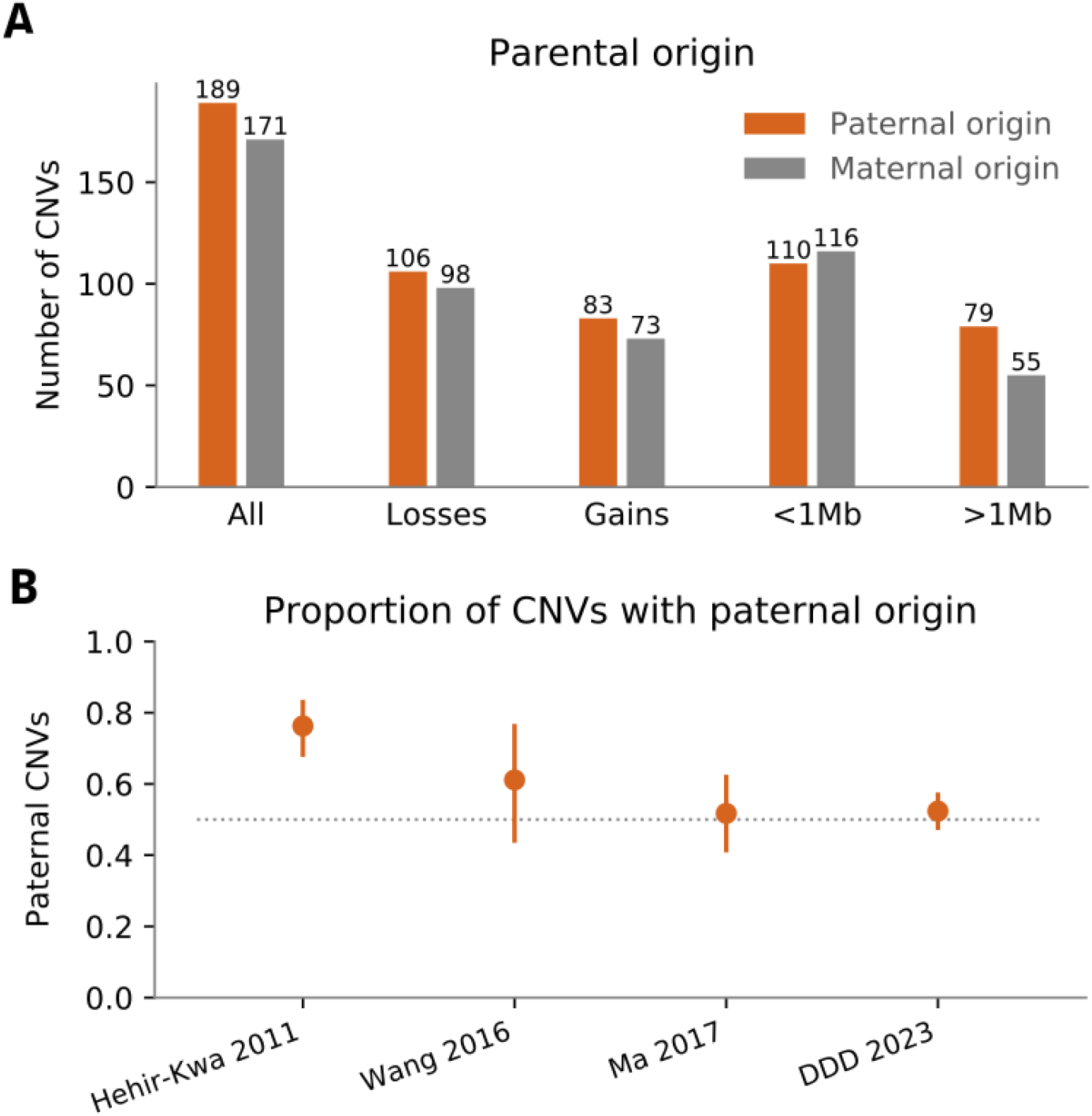
**A**. De novo CNVs identified as part of this study quantified by paternal and maternal origin. None of the categories are significantly enriched for paternal CNVs (Benjamini-Hochberg corrected two-sided binomial tests, p > 0.05). Although we do not find a statistically significant difference between de novo CNVs of paternal or maternal origin (p = 0.37), we observe a higher absolute number of de novo CNVs of paternal origin in our data; however, it is much less prominent than expected from Hehir-Kwa et al.^32^. **B**. Proportion of de novo CNVs with paternal origin in four studies.

Similarly, the mutation rate of *de novo* SNVs is known to increase markedly with paternal age^30^ and more modestly with maternal age^37^. Moreover, the risk of aneuploidies is known to increase with maternal age^38^, but an association between parental age and increased *de novo* CNV has not been established^39^. We did not observe a significant association between *de novo* CNVs and parental age (Supplement S20). While we can be confident that any parental age effect for CNVs must be significantly smaller than for SNVs (p < 1e-270), larger studies are required to determine whether a much more modest parental age effect might exist.

### Pathogenic *de novo* CNVs in DDD ascertained from ES

Of the 598 *de novo* CNVs identified via ES-based calling, we identified 305 (51%) as plausibly pathogenic either due to encompassing many genes as per ACMG and ClinGen guidelines^40^ or due to impacting a known DD-associated gene based on DDG2P^18^ (Supplement S10b). A clinical review of these patients confirmed that these variants were likely to be contributing to the proband’s disorder (Additional File 2), a diagnostic yield of around 3.1%. We note that 86/305 (28%) of these pathogenic CNVs overlapped fewer than three probes in low-resolution 60k CMA and would likely be missed by this assay.

We also examined whether any of the *de novo* CNVs might be contributing to recessive disorders by seeking gene-disrupting variants on the other allele. We identified seven instances of additional truncating variants impacting the same gene as the *de novo* CNV, however all were common in the general population (allele frequency > 0.2) and are thus unlikely to be pathogenic.

With the inclusion of thousands of ultra-conserved non-coding elements in our custom exome sequencing^12^ we also sought noncoding *de novo* CNVs plausibly associated with patient phenotype. In addition to the pathogenic non-coding deletion affecting the 5’ UTR of *ANKRD11* described above, we found three additional patients with a phenotype fully explained by a noncoding deletion: two intersected the promoter of *MEF2C* and are described in more detail elsewhere^41^, and one intersected the promoter of *MBD5*. We also identified an additional patient with a deletion within the promoter of *EHMT1* for which there is currently insufficient evidence to classify as being likely pathogenic or pathogenic.

Prior to recruitment to the DDD study, 7,182 (73%) of the 9,859 participants with trio ES data had previously been clinically tested for large pathogenic CNVs using low-resolution CMA. As such, DDD study participants do not represent an unbiased sample of DD patients, but rather will be depleted of patients with large pathogenic CNVs. Among those who had undergone prior low-resolution CMA testing, we observed 2.6% with a pathogenic CNV identified by one or both of the exon-resolution CMA and ES-based CNV detection. Among the 27% of participants who had not previously received low-resolution CMA, we identified 280 participants (3%) who were likely recruited prior to CMA testing being available in their regional centre. In this group we observed a higher diagnostic yield from *de novo* CNVs of 5.0%. Comparison of the CNV diagnostic yields in these two groups suggests that 52% of pathogenic CNVs detectable from ES are invisible to low resolution arrays (rate ratio test, CI = 31 to 98%, p = 0.044; Figure 5), which is consistent with our simulations (Supplement S8). The CNV diagnostic yield of CMA testing in larger cohorts of patients has been shown in previous studies^4-5^ to be higher, in the range of 10-15%. Therefore our estimate of the added value of CNV detection from ES of 52% is likely to vary depending on the ascertainment of the cohort under study.

**Figure 5:**
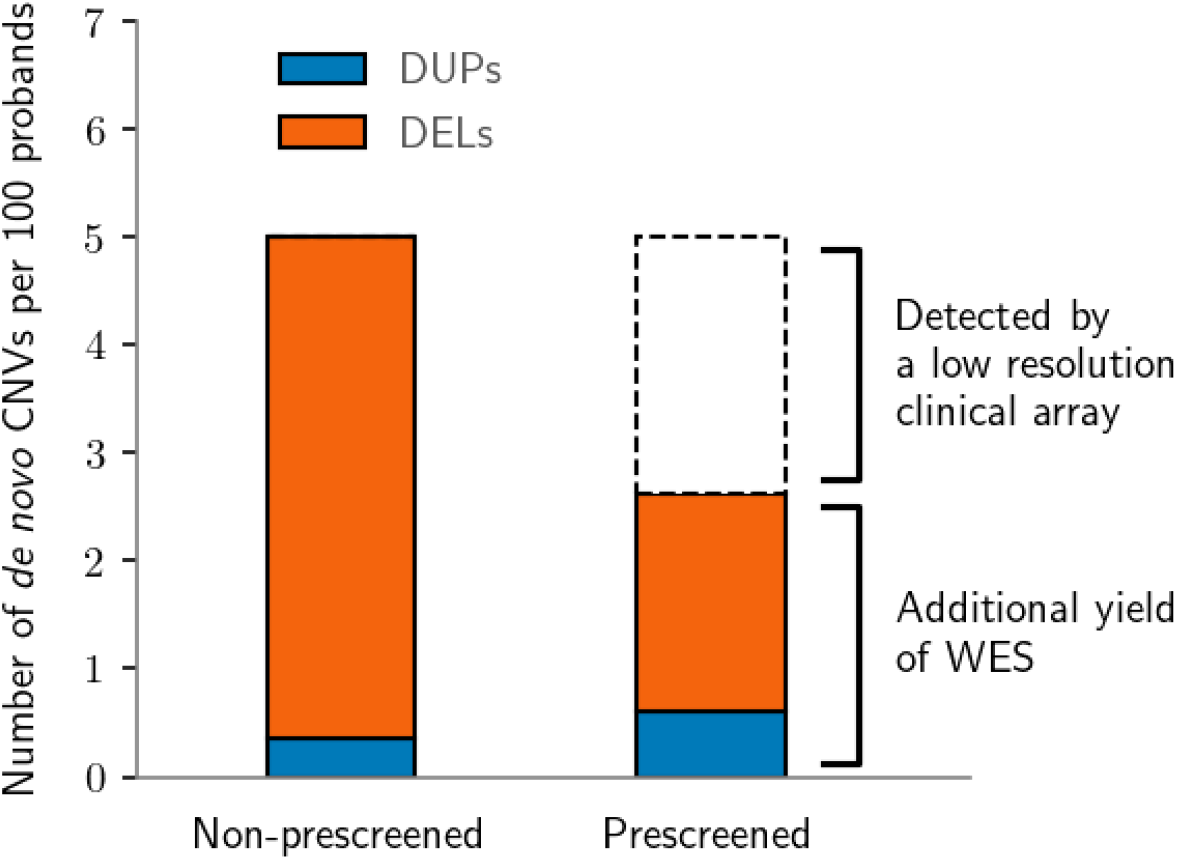
Different discovery rates of pathogenic de novo CNVs were observed in the groups of CMA-untested (14 out of 280; left) and CMA-tested (189 out of 7,182; right) patients.

## Discussion

We developed a CNV calling workflow from ES data that integrates four different calling algorithms using random forest machine learning to generate an ES-based CNV callset of considerably higher quality than achievable with any single calling algorithm. Applying this CNV calling workflow to 9,859 parent-offspring trios participating in the DDD study identified pathogenic CNVs that could not be detected by low resolution CMA, often small CNVs encompassing few exons. Modelling of sensitivity based on probe/bait locations and empirical detection of pathogenic CNVs in a sub-cohort that had not received prior CMA screening, suggests that this ES-based CNV calling workflow likely has high sensitivity to detect the typically large pathogenic CNVs that can be detected by low resolution CMA. Low precision and variable performance of individual ES-based CNV callers was also previously observed in a benchmarking exercise focused on a gold standard dataset^42^, and the value of using machine learning to integrate ES-based CNV calls from different callers was previously validated in a smaller study of 503 patients using an overlapping set of four CNV callers to those used here^43^. However, we note that our random forest model required several iterations of manual curation to optimize and that parameters of the random forest model depend on the noise properties of our data and may need to be re-trained for different data sets.

Comparison of ES-based CNV calling to exon-resolution CMA in 5,197 families suggested that the two approaches have similar sensitivity for pathogenic CNVs. Each approach has incomplete sensitivity to detect CNVs that encompass 1-3 exons, with the result that the combination of the two approaches, while largely concordant, did identify more pathogenic CNVs that would be detected by either approach in isolation. Of DDD probands pre-screened with low resolution CMA, 2.6% had a pathogenic CNV detected by higher resolution assays and the diagnostic yield of pathogenic CNVs in patients that have not previously been screened with low resolution CMA was 5.0%. This is much lower than the >10% diagnostic yield that has been reported in similar patient cohorts, but the difference of 2.4% is comparable to the 1.3% added diagnostic yield reported in a previous, smaller study of patients with neurodevelopmental disorders, the majority of whom had previously been screened with low resolution CMA^44^. We did note that a low proportion of large pathogenic CNVs were hard to call from both exon-resolution assays due to fragmentation into smaller CNV calls by the calling algorithms. This suggests that there are additional improvements to be made in the bioinformatic post-calling merging of CNV calls in order to support robust clinical interpretation with accurate breakpoint definitions with respect to the genes impacted by a CNV.

Overall, we identified 598 *de novo* CNVs in the 9,859 parent-offspring trios of which 305 were clinically interpreted to be contributing to the proband’s clinical phenotype (i.e. classified as Pathogenic or Likely Pathogenic). We did not observe an age or sex bias in the parental origin of these *de novo* CNVs. The lack of sex bias is in contrast to previous, smaller, studies that have suggested a paternal bias for *de novo* CNVs^32,33^. This discordance may be due, in part, to the different size distributions and associated mutational mechanisms being interrogated in the previous studies. Meta-analysing our current data with three previous studies (Supplement S19) does suggest there may be a relatively subtle paternal bias (58%:42%), but larger datasets across the full size range of pathogenic CNVs would help to confirm this.

Population surveys of CNVs across the full size distribution have repeatedly shown that smaller CNVs, below the threshold of detection of low-resolution CMA, are far more numerous and generated at higher mutation rates than larger CNVs. Nonetheless, this study, in combination with previous work, clearly shows that the added diagnostic yield from detecting these smaller CNVs is relatively modest. What matters more in a clinical context is not the total number of CNVs of a given size class, but rather the size distribution of CNVs that disrupt developmentally important genes, which is clearly biased towards very large CNVs that can be detected by low-resolution CMA. In the context of large-scale diagnostic testing of tens of thousands of patients with developmental disorders, ES-based CNV calling is likely to enable a diagnosis in hundreds of families who might well otherwise go undiagnosed. One limitation of our study is that we cannot estimate directly the overall diagnostic yield of ES-based CNV calling as a first-line test due to the prior clinical CMA testing for most of the DDD cohort.

One of the limitations of ES-based CNV calling, as opposed to the custom exon-resolution CMA assay that we used, is the lack of baits to non-coding sequences, meaning that ES-based calling has lower precision in determining the breakpoints of a CNV. This limits the potential for ES-based CNV calling to detect pathogenic CNVs impacting non-coding regulatory elements. In theory, this limitation could be overcome by including a genome-wide ‘backbone’ of non-coding baits in a customised exome design, however, we doubt that, currently, the added diagnostic yield from greater breakpoint precision will be worth the added sequencing costs incurred. Given the high sensitivity of ES-based CNV calling to large pathogenic CNVs detectable by low-resolution CMA, such genome-wide backbone baits are not necessary for ES-based CNV calling to detect these large pathogenic CNVs in the absence of low-resolution CMA. Customising exome designs to include additional baits flanking exons of dosage-sensitive genes to improve sensitivity to detect single exon deletions might be a preferable approach to increase sensitivity of ES-based CNV calling. In principle, increasing the depth of coverage of standard ES should also increase sensitivity for calling single exon CNVs (and for detecting mosaic CNVs).

Our study provides compelling evidence from side-by-side comparison in thousands of families of exon-resolution CMA and ES-based CNV calling that, with appropriate development and deployment of a bioinformatic workflow integrating multiple calling algorithms, ES-based CNV calling has higher sensitivity for pathogenic CNVs than low-resolution CMA and can even render exon-resolution CMA largely redundant. We look forward to similarly scaled side-by-side comparisons of other genomic assays that purport to increase diagnostic yield of pathogenic structural variants (e.g. whole genome sequencing, long read technologies) to accurately quantify the added diagnostic yield over and above the application of best practice bioinformatics pipelines to cheaper assays, enabling diagnostic services to make well-informed cost/benefit decisions.

## Data Availability

Sequence and variant-level data and phenotypic data for the DDD study data are available from the
European Genome-phenome Archive (EGA; https://www.ebi.ac.uk/ega/) with study ID
EGAS00001000775. Clinically interpreted variants and associated phenotypes from the DDD study
are available through DECIPHER (https://decipher.sanger.ac.uk).

## Acknowledgements

We thank the DDD participants and their families – without their trust and confidence this work would not be possible. We thank Dr Nigel P Carter for his pioneering work in the early phase of this study. Parthiban Vijayarangakannan developed the unpublished CoNVex program. We acknowledge the support of the National Institute for Health Research, through the Comprehensive Clinical Research Network. This study makes use of DECIPHER (http://decipher.sanger.ac.uk), which is funded by the Wellcome.The DDD study presents independent research commissioned by the Health Innovation Challenge Fund [grant number HICF-1009-003], a parallel funding partnership between Wellcome and the Department of Health, and the Wellcome Sanger Institute [grant number WT098051]. For the purpose of open access, the author has applied a CC BY public copyright licence to any Author Accepted Manuscript version arising from this submission. The views expressed in this publication are those of the author(s) and not necessarily those of Wellcome or the Department of Health.

## Author Contributions

T.W.F., G.G., R.Y.E. and P.D. performed CNV calling; P.D. processed and analyzed the data with contributions from E.J.G.; P.D., E.J.G. and J.K. performed statistical analyses; H.V.F. performed clinical review; E.J.G., P.D. and M.E.H. wrote the manuscript; C.F.W, H.V.F and M.E.H oversaw the project.

## Ethics Statement

M.E.H. is a co-founder of, consultant to, and holds shares in, Congenica Ltd, a genetics diagnostic company. E.J.G. is an employee of and holds shares in Adrestia Therapeutics Ltd. Informed and written consent was obtained for all families and the study was approved by the UK Research Ethics Committee (10/H0305/83, granted by the Cambridge South REC, and GEN/284/12 granted by the Republic of Ireland REC).

