## Supplementary File for "Detection and characterisation of copy number variants from exome sequencing in the DDD study"

#### S1 CNV calling from aCGH data

The custom CGH microarray used for CNV discovery was designed to have good power to detect single exon CNVs, by using 5 probes per exon, as well as having a dense backbone of  700k probes outside of exons to facilitate calling of large CNVs and definition of CNV breakpoints. The exon-CGH array is composed of two 1 million probe Agilent arrays and has been heavily targeted towards genes and ultra-conserved elements throughout the human genome. The entire set of GENCODE genes, along with regulatory and mRNA coding elements have been tiled, using a minimum of five oligo-nucleotide probes per exon. The remainder of the probes on the array constitute a high-resolution backbone with a median probe spacing of 5 kb.

The aCGH CNV calls were obtained by CNsolidate (**ref**) which incorporates 12 independent change point detection algorithms and makes use of an expert voting system, 6 of the component algorithms have been described previously[1, 2, 3, 4, 5, 6] and 6 were developed in house. Using this approach, CNsolidate is able to rank detections based on differential weighting functions between component voter algorithms. CNsolidate derives all of its parameter definitions and weighting functions using a number of predictive variables drawn from the input data characteristic. Accurately ranking detections across data sets of variable qualities allows fine gain control over the the balance between type 1 and type 2 errors using a single threshold.

#### S2 CNV calling from ES data

The ES data were generated using one of two custom bait designs based on the Agilent SureSelect v3 and v5 exomes, supplemented with additional probes targeting 6,000 high value non-coding regions comprising 12.9% of the total sequence[7]. The sequencing was performed as described previously in [8]. The paired-end 75 bp sequencing reads were aligned onto the GRCh37d5 reference genome with bwa[9]. Four programs were then used to generate the initial raw callset: CANOES[10], CLAMMS[11], CoNVex[12], and XHMM[13]; see Supplement S3 and S4 for more details. Conflicting breakpoints of calls generated by these four programs were resolved by splitting overlapping regions into segments and choosing the most likely state (deletion, duplication, or normal) for each candidate CNV according to a simple probabilistic model based on the ratio of observed and expected read depth (log2 ratio - L2R) (Supplement S21). The following metrics were used in the random forest classification: concordance (the number of callers that made the call); qualities assigned by the callers (or 0 if the call was not made by a caller); read depth fraction (the ratio of the mean depth in the target and in the flanking regions); number of probes in the call; number of probes in the flanking regions; the length of the call; GC content; the mappability; the mean L2R value; the standard deviation of L2R values; the difference in the mean L2R values between the target and the flanking regions; and the L2R p-value (Supplement S21). The random forest classifier was trained on a manually curated subset of calls, separately for deletions and duplications, and was run with 150 trees in the forest. It achieved the training accuracy of 95% for deletions and 96% for duplications, as estimated by out-of-bag sampling. The most important classification annotations were the concordance between callers and L2R-based metrics for duplications (Supplement S23/A), and L2R-based metrics with the quality scores of CANOES and CoNVex for deletions (Supplement S23/B). The best scoring candidate calls were then manually inspected, the training set extended, and the model retrained and reapplied.

CNV calling was performed on a population level with all samples included and both probands and their parents analysed equally. A two-step procedure was then used to determine *de novo* CNVs from this initial callset. First, raw parental calls made by all programs were used to exclude overlapping calls. Second, the more prohibitive parental version of the simple L2R model (dashed lines in Supplement S21) was used to exclude any potential inherited CNVs that might have been missed by the callers.

The callset was then filtered at a strict random forest threshold of 0.95 which was chosen based on manual inspection of quality score distributions of curated true and false calls. Further we excluded calls present in known variable regions, such as immunoglobulin and T-cell receptor genes (Supplement S22), and CNVs observed in unrelated unaffected parents. In both cases a 75% reciprocal overlap was required to exclude the call. The sensitivity of the callsets was evaluated using a truth set of 280 clinically validated CNVs which were discovered previously using multiple platforms. The number of apparent *de novo* CNVs per-sample was used as a proxy for specificity.

#### S3 Programs and their versions

| | | |
|---|---|---|
| Short reads alignment | | |
| `bwa` | `0.5.10` | `bwa aln -q 15`<br>`bwa sampe` |
| CNV calling | | |
| `CANOES` | `Jan-04-2019` | `http://www.columbia.edu/ ys2411/canoes` |
| `CLAMMS` | `f486d6b` | `https://github.com/rgcgithub/clamms.git` |
| `CoNVex` | `Jan-05-2017` | `ftp://ftp.sanger.ac.uk/pub/users/pv1/CoNVex` |
| `XHMM` | `cc14e52` | `https://bitbucket.org/statgen/xhmm/src/master` |
| CNV analysis | | |
| `annot-regs` | `69dfbd2` | `https://github.com/pd3/utils` |
| `perm-test` | `69dfbd2` | `https://github.com/pd3/utils` |
| `recurrence-test` | `9abe601` | `https://github.com/pd3/utils` |
| `bcftools/parental-origin` | `4caf1fd` | `https://github.com/samtools/bcftools` |
| Scripts and pipelines used in this work | | |
| | | `https://github.com/pd3/cnv-paper-2020.git` |

#### S4 CoNVex algorithm

The CoNVex algorithm utilises the read depth information in probe regions, compares it to a reference depth profile and detects copy number variable segments using an error-weighted score and the Smith-Waterman algorithm applied on log2 ratio of depth over median depth.

##### Estimation of read depth

Exome capture regions were extended by 100bp on either side and overlapping regions merged. The mean depth across all bases was then calculated in every region, with duplicate reads removed.

##### Calculation of $\log_2$ ratio and error correction

Regions of varying copy number may have high or low depth, but majority of regions are copy number neutral and form the median of the distribution. Therefore, CoNVex uses the median depth as a baseline of sequencing coverage. A correlation matrix of median depths is built and for each sample a subset of 25 most similar reference samples is selected. Then in every bait region it calculates the $\log_2$ ratio of the observed depth over the median depth of reference samples.

Systematic erros may occur at the hybridization level as well as at the sequencing level, and read depth is known to be influenced by GC content. A generalized additive model (GAM) with multivariate cubic splines is used to correct systematic errors in $\log_2$ ratio for each probe region, hybridization free energy $\Delta G$ and melting temperature $T_m$ are the covariates.

Variation in the $\log_2$ ratio may prevail even after the correction of systematic errors. Bait regions are classified into bins based on the number of reads and the median absolute deviation (MAD) of the $\log_2$ ratio within each bin is calculated as a measure of noise. A heuristic score is then calculated using the corrected $\log_2$ ratio and the bin-wise MAD of the $\log_2$ ratio. These scores are then error-weighted and used further for the change point detection.

##### Change point detection

The Smith-Waterman algorithm is used for the detection of change points from the error-weighted heuristic scores to efficiently detect and merge the copy number varying, high (duplication) or low (deletions) scoring bait regions. This is a local implementation of the algorithm based on SW-array, a similar algorithm used for the aCGH data.

CNV detection starts with the selection of calls generated by the Smith-Waterman algorithm. For each CNV call, an SW-score is calculated based on the heuristic scores of the number of merged bait regions. This score is adjusted for the number of merged regions by dividing the score with the square root of the number of bait regions. The final score is the adjusted SW-score and named as CoNVex score. A cut-off has been applied based on our specificity analysis that compares the proportion of known (with a population frequency of 1% or more) CNVs vs the CoNVex score.

##### Implementation and availability

CoNVex is written R and java, the whole package is available from the WTSI public FTP repository `ftp://ftp.sanger.ac.uk/pub/users/pv1/CoNVex`.

##### Author of the program

Parthiban Vijayarangakannan

#### S5 Diagnostic DDD Pipeline

CoNVex was run on all proband and parent samples, and inheritance predicted with CIFER (`https://github.com/jeremymcrae/cifer`), aCGH data were available only for probands. CoNVex and aCGH calls were then merged and annotated with overlap with rare and common CNVs and internal frequency within the DDD samples.

Separate aCGH and WES filters were then applied on CNsolidate and CoNVex calls. If a CNV was detected by both aCGH and WES, the aCGH filters were used. CNVs was not submitted for a clinical review if any of the following were true:

aCGH filters:

- $W$ score $< 0.45$
- Call quality $p$ value $> 0.01$
- Common forwards $> 0.8$
- Duplications - mean log2ratio $< 0.4$
- Deletions - mean log2ratio $> -0.5$
- abs(log2ratio/MAD) $< 10$
- No overlapping exons
- Internal frequency $> 0.01$
- CIFER inheritance = false positive

WES filters:

- CoNVex score $\leq 7$
- Internal frequency $> 0.01$
- No overlapping exons
- abs(log2ratio/MAD) $< 10$
- Duplications - mean log2ratio $< 0.4$
- Deletions - mean log2ratio $> -0.5$
- Common forwards $> 0.8$

In addition, on the X chromosome the mean log2ratio is summed for all probes for each individual. If the sum of the log2ratio on X for that individual is $< -5000$ or $> 7000$ then all calls on X are discarded for this person.

Additional filters for deletions - deletions which are predicted to be not inherited or uncertain by CIFER fail if two or more of the following are true:

- mean log2ratio $< -1.5$
- convex score $< 15$
- MAD $> 0.15$

If the filters are passed and a CNV is $> 1,000,000$ bases then it is reported. If the CNV is $\leq 1,000,000$ bases then it must overlap a gene in DDG2P and the inheritance seen for this CNV must be consistent with what is known for the gene. The filtering is under constant revision and improvement.

The package for clinical filtering is available from
`https://github.com/jeremymcrae/clinical-filter/tree/master/clinicalfilter`.

#### S6 Gene Enrichment Testing

To test enrichment of CNVs in genes associated with developmental disorders in contrast to all genes (Figure 2), we compiled a subset of 800 genes in which a *de novo* mutation is sufficient to cause a disease (i.e. chromosome X-linked, hemizygous and monoallelic genes) from the Developmental Disorders Genotype-To-Phenotype Database (DDG2P, `https://www.ebi.ac.uk/gene2phenotype/downloads/DDG2P.csv.gz` version 2022-10-17; [14]) restricted to categories "definitive" or "strong" and in further text referred to simply as the "DN-DD genes".

We performed a permutation test in which calls were randomly placed along the genome and the number of placements that overlapped a coding sequence of a DN-DD gene was collected. The test was run separately for duplications and deletions, each consisted of 1e10 permutations.

Similarly for gene recurrence tests (Figure 3b-c), calls were randomly placed on the genome and in each iteration we counted how many times a gene was hit by fewer calls than observed. The test was run separately for duplications and deletions, each consisted of 5e9 permutations.

For efficiency CNVs bigger than 10Mbp were excluded because large aberrations are likely to affect genes in both categories equally, and in random placements we considered only exome bait regions accessible to the sequencing experiment. Only autosomal chromosomes were considered in the simulations. The tests were implemented in the programs perm-test and recurrence-test (Supplement S3).

#### S7 Comparison Between Platforms and Callsets

Based on the premise that CNV calling sensitivity is directly correlated with the number of probes or baits intersecting a CNV, the expected sensitivity of various platforms (60k, 180k, 2M aCGH and ES) was estimated by randomly placing 6,412 CNVs with a variant size distribution derived from 1000 Genomes Project SV data(25) (Supplement S8) and evaluating the number of probes affected across 500,000 simulated runs. The GRCh37.87 gene model (`ftp://ftp.ensembl.org/pub/grch37/release-87/gff3/homo_sapiens/Homo_sapiens.GRCh37.87.gff3.gz`) was used to determine the coding regions of the genes, with overlapping exons merged and counted as a single exon. Chromosome X was included in the simulation.

When comparing the callsets we ensured that the same set of samples were considered and that appropriate thresholds were used to account for different specificity and sensitivity profiles. Therefore, in order to compare the sensitivity of aCGH and ES, the default recommended CNsolidate cutoff 0.5 was used to filter the aCGH calls and the *de novo* mutation rate estimated at this threshold was then used to determine the corresponding cutoff with the same specificity in ES data. In order to account for different probe density and platform design, an overlap of arbitrary size was sufficient to deem two calls as matching.

All comparisons relied on the program annot-regs (Supplement S3).

The assays used in the comparison were

**60k aCGH**
: A-MEXP-2268, Agilent CGH ISCA v2

**180k aCGH**
: ACGH OGT CytoSure Constitutional v3

**2M aCGH**
: two custom 1 million probe Agilent arrays designed to target genes and ultra-conserved elements throughout the human genome. The entire set of Gencode genes (version 17), along with some high value regulatory and mRNA coding elements have been tiled, using a minimum of 5 oligo-nucleotide probes per exon. Additionally the array maintains the presence of a high-resolution backbone with a median probe spacing of 2Kb (Amadid No.s 031220/031221). For details see Additional File 1.

**ES**
: custom bait designs based on the Agilent SureSelect v3 and v5 exomes, supplemented with additional probes targeting ∼6,000 high value non-coding regions comprising 12.9% of the total targeted sequence

### S8 Predicted sensitivity of aCGH and WES

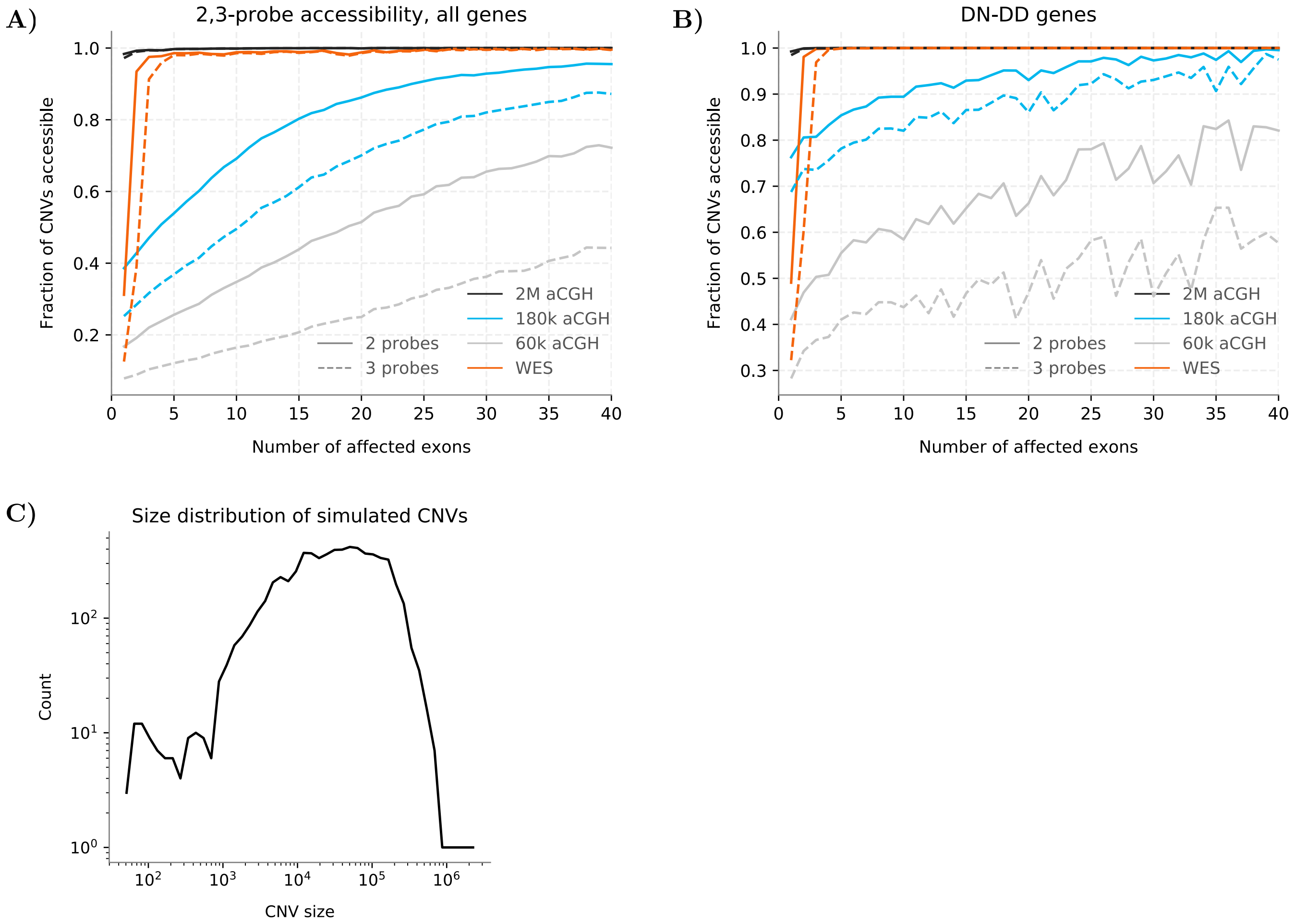


Figure S8: The predicted sensitivity of whole-exome sequencing (WES; orange lines), custom design CGH array with 2M probes (black), and two typical CGH arrays in clinical use with 180k probes (OGT's CytoSure Constitutional v3; blue) and 60k probes (A-MEXP-2268, Agilent CGH ISCA v2; grey) in all genes (**A**) and DN-DD genes (**B**). The accessibility estimate is based on a simple discovery model which requires a minimum of 2 (solid lines) or 3 (dashed lines) affected probes for a successful detection. (**C**) The simulation used CNVs identified by the 1000 Genomes Project (Sudmant et al. 2015), restricted to bait regions used by the DDD project (see main text Methods).

#### S9 Counts of raw aCGH and WES calls

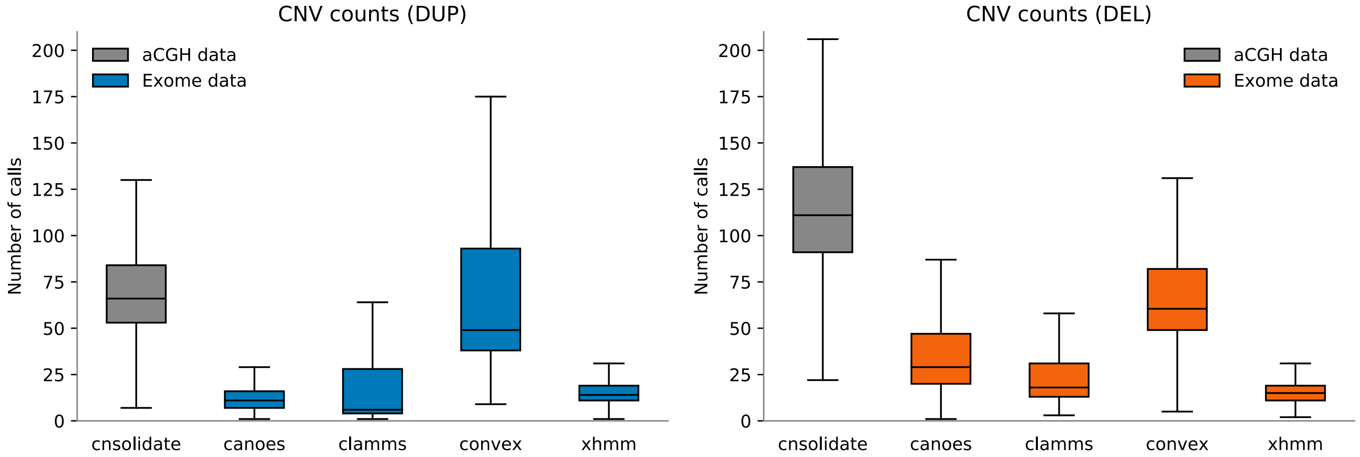


Figure S9: The per-sample number of raw, unfiltered calls as generated by CNsolidate from 2M aCGH data (grey) and by CANOES, CLAMMS, CoNVex, and XHMM (colored) from WES data, prior to assigning inheritance status. There were large inter- and intra-platform differences, for both duplications (left) and deletions (right).

#### S10 CNV calling and filtering of ES calls

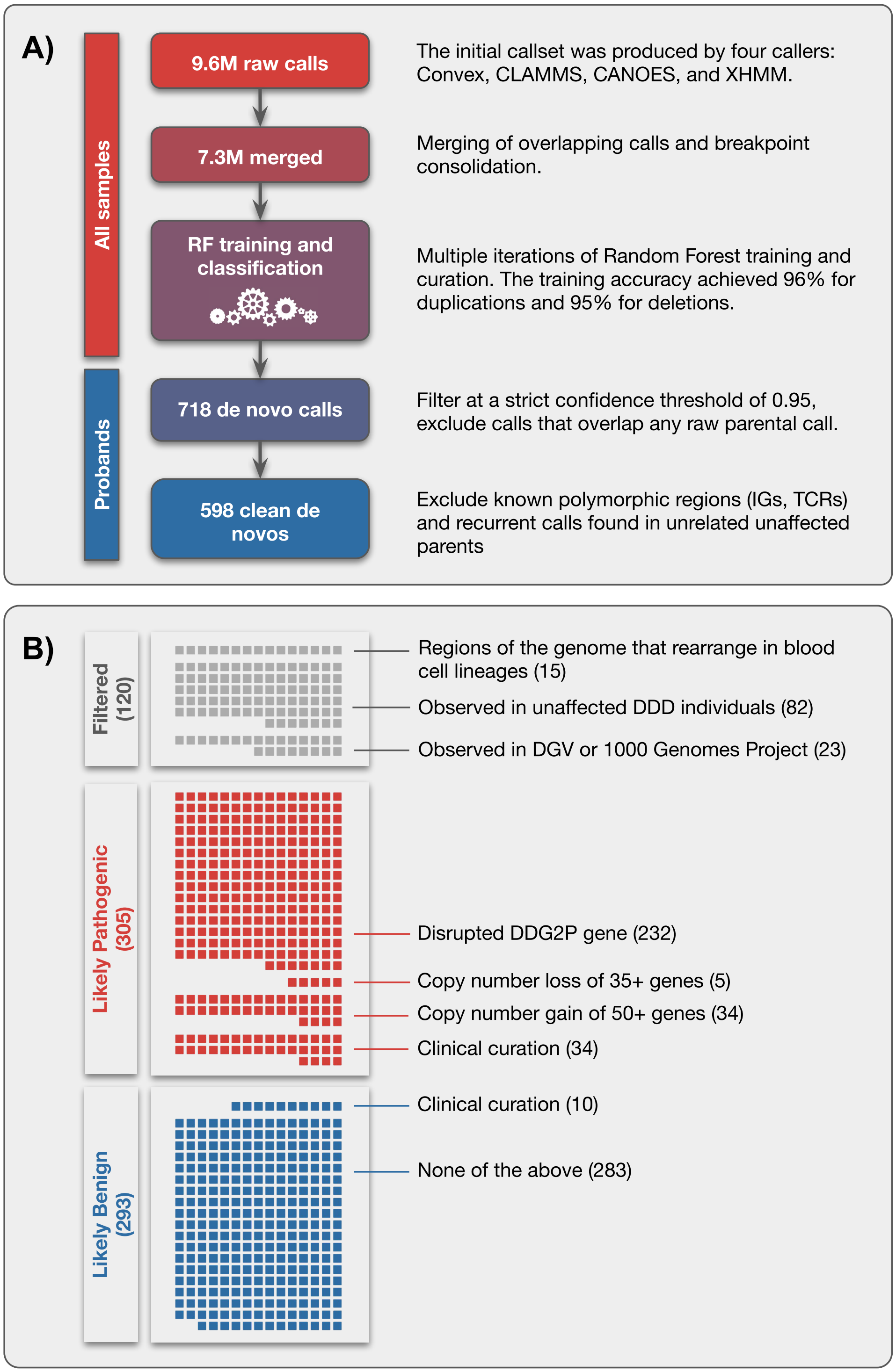


Figure S10: Outline of the CNV calling and filtering pipeline in the DDD study from ES data (**A**) and classification of the 718 *de novo* CNVs to likely pathogenic and benign CNVs (**B**).

#### S11 Distribution of random forest scores

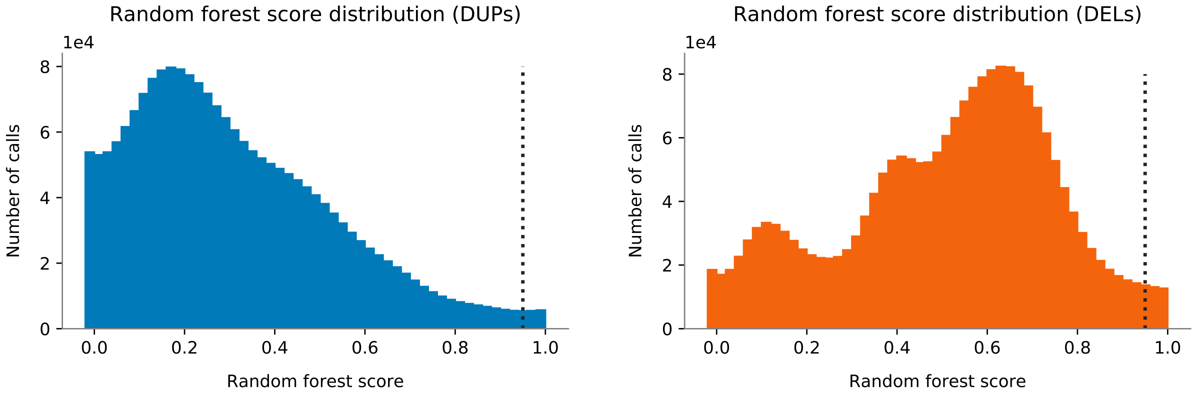


Figure S11: The distribution of random forest classification scores for duplications (left) and deletions (right). The dotted line indicates the filtering threshold 0.95.

#### S12 Comparison of CNVs missed by 2M aCGH and WES

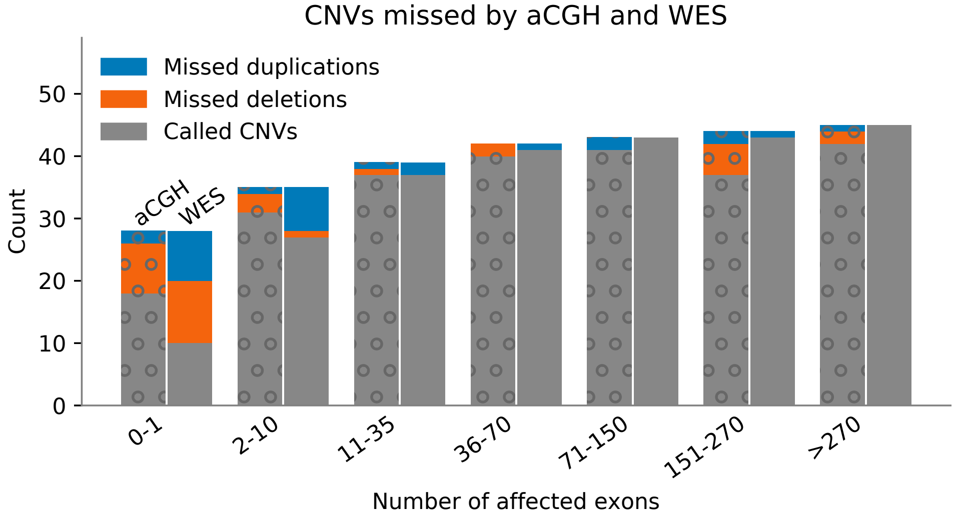


Figure S12: Sensitivity of aCGH and WES-based approaches to a truth-set of 276 CNVs as a factor of number of affected exons (x-axis). The default recommended CNsolidate cutoff 0.5 was used to filter the aCGH calls and the *de novo* mutation rate at this threshold was then used to determine the corresponding cutoffs with the same specificity in WES data (0.78 for duplications and 0.63 for deletions).

#### S13 CNVs affecting multiple vs single gene

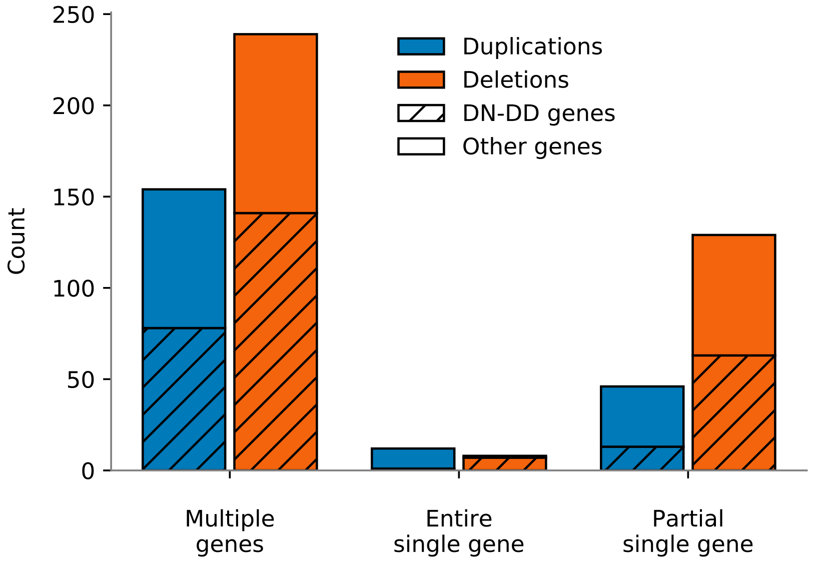


Figure S13: The number CNVs affecting coding sequence of multiple genes or of a single gene, partially or the entire gene.

#### S14 Permutation tests

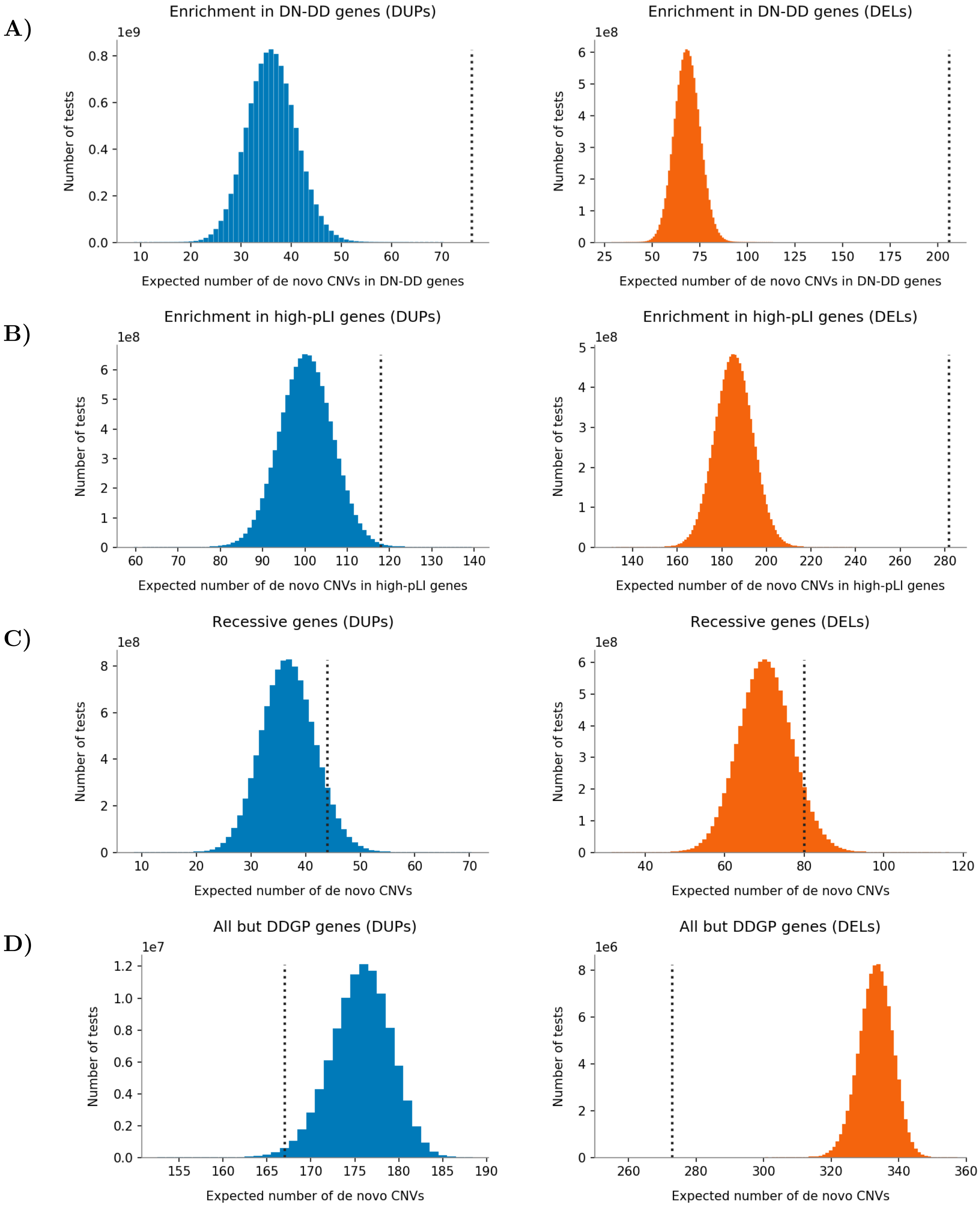


Figure S14: Expected number of duplications and deletions overlapping (**A**) a DN-DD gene, (**B**) a constrained gene with pLI $> 0.9$, (**C**) a recessive DD gene not constrained for deleterious genetic variation (pLI $< 0.9$), (**D**) a gene not listed in DDGP. The distributions show the number of CNVs overlapping the set of target genes in a permutation test which consisted of 1e10 iterations each. The dotted line shows the actual number of duplications and deletions observed in our data set.

#### S15 Entire vs partial CNVs

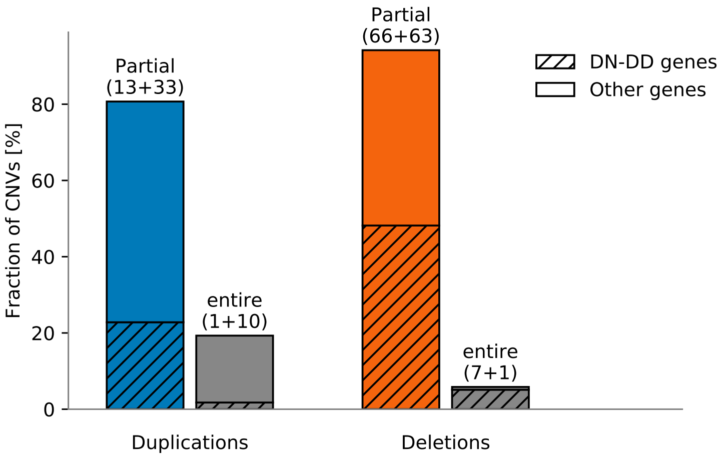


Figure S15: The number of single gene duplications and deletions affecting the gene either partialy or in its entirety. The numbers indicate the counts affecting coding sequence of DN-DD and non DN-DD genes, respectively.

#### S16 Genes passing genome-wide significance threshold

Table S16a: List of ten DN-DD genes that pass genome-wide significance threshold.

*ANKRD11*
*EFTUD2*
*ELN*
*FBXO11*
*KIF22*
*LZTR1*
*MAPK1*
*MED13L*
*PRRT2*
*TBX1*

Table S16b: List of 9 recurrently mutated loci known to be associated with DD that have reached genome-wide significance threshold, sorted by the number of *de novo* CNVs observed in our study.

| | | | |
|---|---|---|---|
| 22q11 | 22 : 19,040,000 - 22,470,000 | 61 | OMIM; McDonald-McGinn *et al*, (PMID:27189754 ) |
| 16p11.2 | 16 : 29,650,000 - 31,200,000 | 35 | Shinawi *et al*, 2009 (PMID:19914906) |
| 16p11.2 | 16 : 28,820,000 - 30,050,000 | 25 | Bachmann-Gagescu *et al*, 2010 (PMID:20808231) |
| 7q11.23 | 7 : 72,740,000 - 75,140,000 | 18 | Morris *et al*, 2015 (PMID:26333794) |
| 15q13.3 | 15 : 29,160,000 - 33,460,000 | 11 | Rosenfeld *et al*, 2011 (PMID:21248749) |
| 15q13.3 | 15 : 31,080,000 - 33,460,000 | 11 | Deutsch *et al* 2016 (PMID:26257138) |
| 15q13.3 | 15 : 32,020,000 - 33,460,000 | 9 | Gillentine & Schaaf, 2015 (PMID:26095975) |
| 16p12.2 | 16 : 21,950,000 - 23,430,000 | 2 | Girirajan *et al*, 2010 (PMID:20154674) |

#### S17 Parental Contribution of CNVs

In order to determine whether a *de novo* CNV originated on the maternal or the paternal allele, we used informative SNVs called from ES data in the affected region. It is straightforward to determine the parental origin for deletions by comparing the combination of parental genotypes with homozygous SNVs in the child. For duplications, the fraction of alternate and reference reads observed at heterozygous SNVs in the child was used to model the probability of the duplicated allele originating on the paternal or the maternal chromosomal copy (Supplement S18). The method was implemented in `bcftools` plugin `parental-origin` and thoroughly verified by another implementation (`https://github.com/pd3/cnv-paper-2020`) and an independent manual validation, including a sample identity and sex verification to exclude potential sample swaps.

Further, to test the association between parental age and risk for *de novo* CNVs, we used a generalized linear regression with a Poisson error distribution and the link function set to "identity". The response variable was the total number of *de novo* CNVs per proband and the maternal and paternal ages independent covariates:

$$N_{CNV} \approx \text{age}_{paternal} + \text{age}_{maternal}$$

We then used the t-statistic to test whether the age coefficients, which represent the change in mutation rate of *de novo* CNVs with the age, are different for SNVs. For plotting, the probands were divided by age into nine quantile groups with median age representing the group's age.

Only autosomal chromosomes were considered in the analyses.

#### S18 Parental origin ascertainment

In order to determine whether a CNV originated on the paternal or maternal chromosome, we implemented the `bcftools` plugin `parental-origin`. The logic of the program is schematically represented in Figure S18 and its probability model explained below.

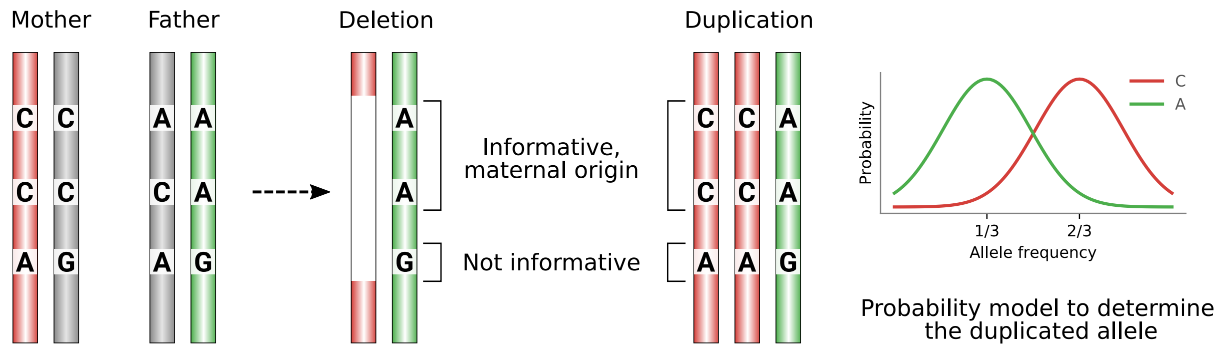


Figure S18: Parental origin of deletions and duplications can be determined from informative SNPs.

##### Deletions

Let $d \in \{0, 1, 2\}$ denote the alternate allele dosage, $G \in \{P, M, F\}$ the proband, mother and father, and $G_d$ all possible genotype likelihoods for the proband, mother and father. For example, $P_0$ denotes the likelihood of the homozygous reference genotype in the proband, $M_1$ is the likelihood of the heterozygous genotype in the mother, and $F_2$ the likelihood of the homozygous alternate genotype in the father. These values are obtained from the FORMAT/PL field of a trio VCF file.

The probability of a deleted paternal allele $P_i(F)$ at site $i$ can be then expressed as

$$
\begin{aligned}
P_i(F) = P_0 &\left[\frac{1}{2}M_0F_0 + \frac{2}{3}M_0F_1 + M_0F_2 + \frac{1}{3}M_1F_0 + \frac{1}{2}M_1F_1 + M_1F_2\right] \\
+ P_2 &\left[\frac{1}{2}M_2F_2 + \frac{2}{3}M_2F_1 + M_2F_0 + \frac{1}{3}M_1F_2 + \frac{1}{2}M_1F_1 + M_1F_0\right]
\end{aligned}
$$

and the probability of maternal origin $P_i(M)$ as

$$
\begin{aligned}
P_i(M) = P_0 &\left[\frac{1}{2}M_0F_0 + \frac{2}{3}M_1F_0 + M_2F_0 + \frac{1}{3}M_0F_1 + \frac{1}{2}M_1F_1 + M_2F_1\right] \\
+ P_2 &\left[\frac{1}{2}M_2F_2 + \frac{2}{3}M_1F_2 + M_0F_2 + \frac{1}{3}M_2F_1 + \frac{1}{2}M_1F_1 + M_0F_1\right].
\end{aligned}
$$

Then we take the product of site probabilities and determine the parental origin as the more likely, assigning the call quality

$$
Q = \left|\log_{10}\prod P_i(M) - \log_{10}\prod P_i(F)\right|.
$$

##### Duplications

For duplications we need to introduce new terms $P_{rra}$ and $P_{raa}$ to express the probability of an increased copy number state in the proband. Let again $P_1$ be the likelihood of the heterozygous genotype in the proband and $B$ a one-sided binomial test of the deviation from the expected ratio of reference and alternate alelle (1/3 or 2/3), $N_r$ the number of observed reads with the reference allele, and $N_a$ the number of reads with the alternate allele, then

$$
\begin{aligned}
P_{rra} &= P_1\, B(X \geq N_r; N_a, \mu = 1/3), \\
P_{raa} &= P_1\, B(X \leq N_r; N_a, \mu = 2/3).
\end{aligned}
$$

Then the probability of a single copy gain can be expressed as

$$
\begin{aligned}
P_i(F) &= P_{rra}\left[M_1F_0 + M_2F_0 + \frac{1}{2}M_1F_1 + M_2F_1\right] \\
&+ P_{raa}\left[M_1F_2 + M_0F_2 + \frac{1}{2}M_1F_1 + M_0F_1\right], \\
P_i(M) &= P_{rra}\left[M_0F_1 + M_0F_2 + \frac{1}{2}M_1F_1 + M_1F_2\right] \\
&+ P_{raa}\left[M_2F_1 + M_2F_0 + \frac{1}{2}M_1F_1 + M_1F_0\right],
\end{aligned}
$$

with total probabilities and the call quality calculated the same way as for deletions.

#### S19 Parental origin comparison

| Study | Paternal origin [%] | N (paternal, maternal) | P(binom) |
|---|---|---|---|
| Hehir-Kwa *et al.* 2011 | 76.3 | 118 (90, 28) | 8.9e-9 |
| Wang *et al.* 2016 | 61.1 | 36 (22, 14) | 0.24 |
| Ma *et al.* 2017 | 51.7 | 87 (45, 42) | 0.83 |
| This study | 52.5 | 360 (189, 171) | 0.37 |
| Combined | 57.6 | 601 (346, 255) | 2.4e-4 |

The bias in paternal origin (or the lack of) as observed across four studies. All but one were underpowered for the observed effect and the bias was less prominent in later studies.

#### S20 CNV rate vs parental age

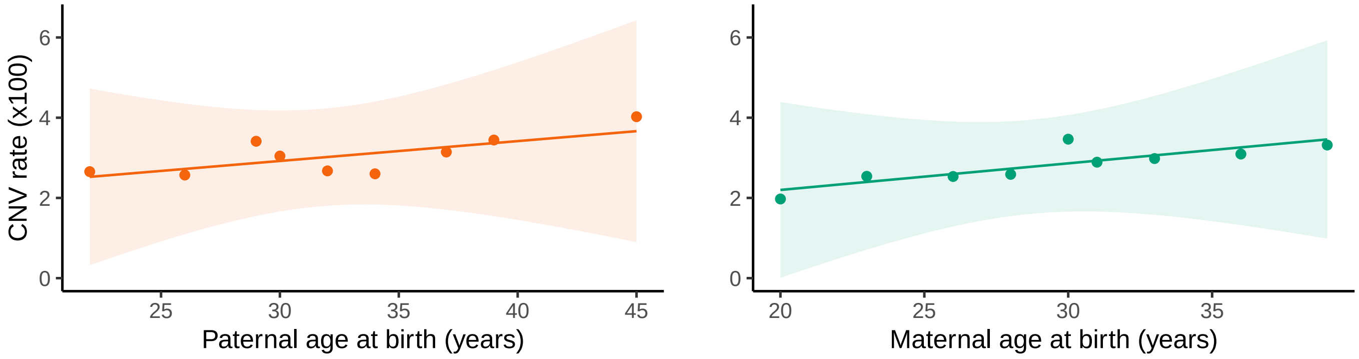


Figure S20: The change of *de novo* CNV formation rate with age is not significantly different from no increase in rate at all (paternal $p = 0.58$, maternal $p = 0.09$). We note that our tests have low power for the observed increase of 0.01 *de novo* CNVs per paternal age holding maternal age constant and 0.04 CNVs per maternal age.

#### S21 Determining the most likely CNV state

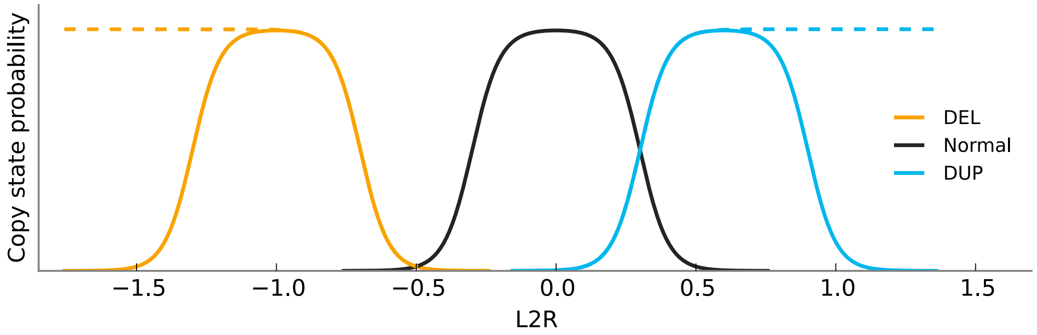


Figure S21: A simple model used to consolidate conflicting breakpoints of calls made by multiple callers. The log ratio of expected and observed normalized depth (L2R) should be centered at -1.0 for one-copy loss, 0 for normal states, and 0.5 for one-copy gains. The model takes into account noise observed in real data. The dashed lines indicate a more permissive model which was applied on parental data to exclude inherited variants and generate a high-confidence *de novo* set.

#### S22 Excluded variable regions

| | | | |
|---|---|---|---|
| chr2 | 88937989 | 90000000 | Ig kappa |
| chr7 | 38240000 | 38380000 | T-cell receptor (TCR) gamma |
| chr7 | 141998851 | 142510972 | TCR beta |
| chr14 | 21150000 | 22100000 | TCR alpha and delta |
| chr14 | 106064028 | 106330469 | Ig heavy chain |
| chr22 | 22380474 | 23265085 | Ig lambda light chain |

#### S23 Metrics used in random forest filtering

A)

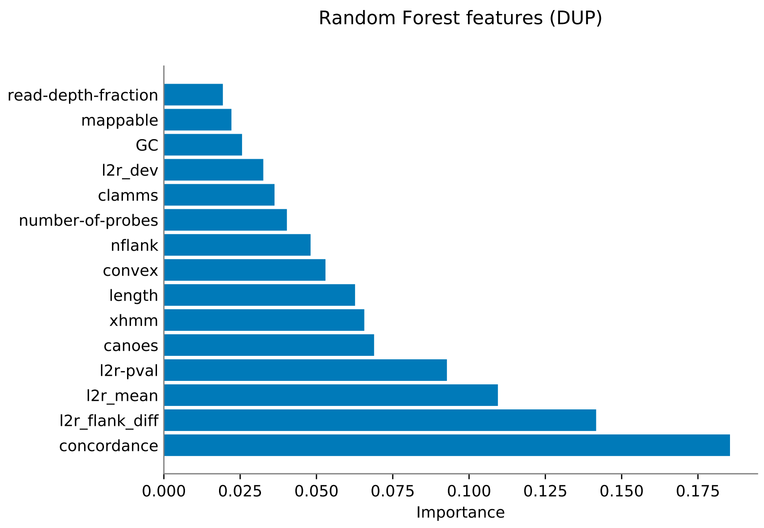


B)

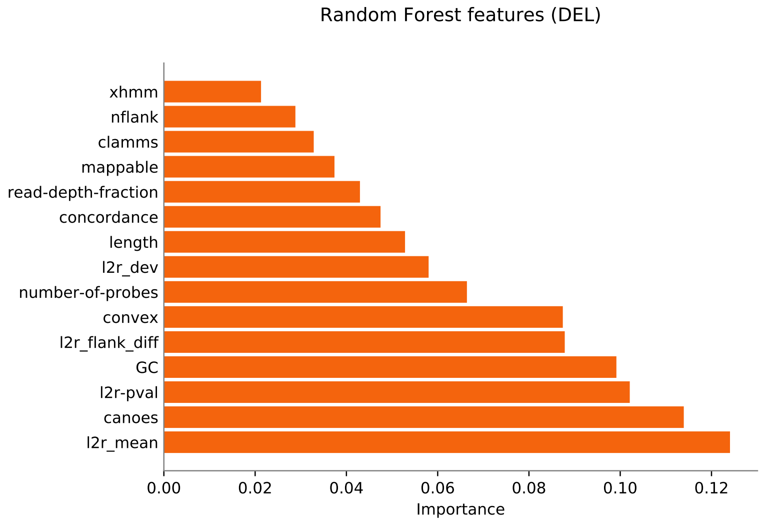


Figure S23: The performance of metrics used in random forest classification. For duplications (**A**), the most predictive was the concordance between callers (how many programs made the call) and L2R associated metrics. For deletions (**B**), most predictive were the L2R metrics, GC content, and the CANOES and CoNVex qualities.

#### S24 Distribution of quality scores

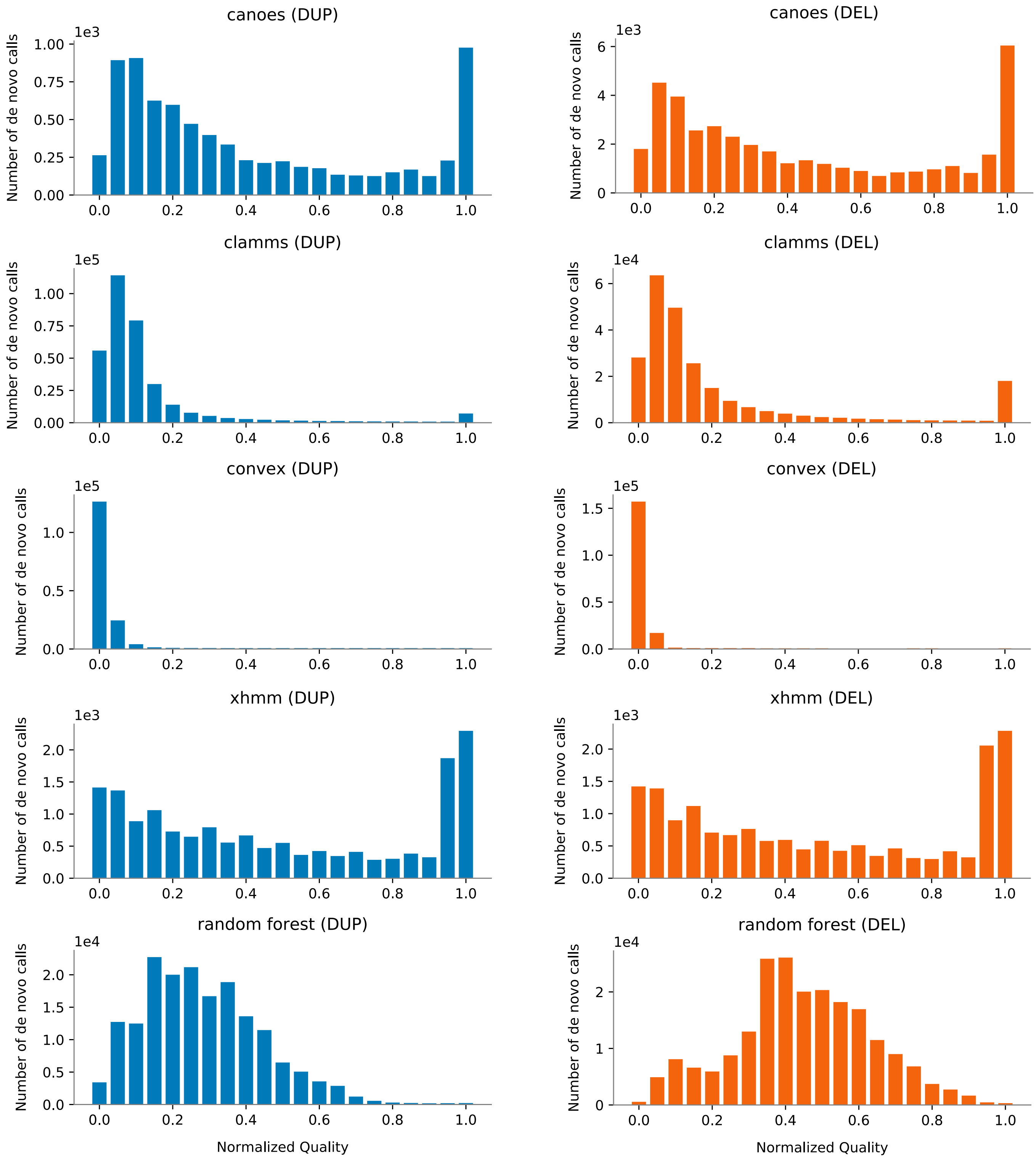


Figure S24: The distribution of quality scores of candidate *de novo* calls produced by CANOES, CLAMMS, CoNVex, XHMM and random forest classification.
