## Additional File 1 for "Detection and characterisation of copy number variants from exome sequencing in the DDD study"

### Design of the DDD array-CGH microarray

**Objectives:**

1. To identify novel pathogenic copy number variants in children with developmental disorders

2. To enable the design of an optimal, smaller array for diagnostic applications

3. To identify whether some common CNVs might be modifiers of more penetrant alleles

#### Design parameters:

- 2x Agilent 1M array format, with probes allocated to arrays so as not to split any individual chromosomal arms across arrays
- 5 probes per targeted locus, to allow deletion/duplication of a single functional locus to be detected
- Probes prioritised on the basis of Agilent *in silico* probe performance metric, with a minimum probe score of 0.4
- Backbone probes chosen to minimize large gaps
- Control probes in CNVs well-tagged by SNPs to allow sample tracking between aCGH arrays and with SNP data
- Set of CNV genotyping probes for CNVs un-tagged by SNPs (n=1662 CNVs)
- Retain Agilent normalization and control group probes per array (N=16488 on each array)
- Probes mapping to a chromosome are retained on the same array

#### Targeted Loci:

##### *Functional sequences:*

***Rationale:*** *to enrich for probes in known or putative functional sequences to maximize sensitivity to detect pathogenic deletions/duplications*

**Genes**
*Definition:* GENCODE annotation levels 1,2 and 3 on HG19, downloaded on 22$^{nd}$ July 2010, including exons in protein coding transcripts and miRNAs (merged into non-redundant set)

**Putative regulatory elements – annotated enhancers**
*Definition:* proven *in vivo* activity in mouse embryos from the Enhancer DB site, downloaded on 3$^{rd}$ August 2010. in HG19 coordinates

**Putative regulatory elements – highly conserved non-coding sequences**
*Definition:* PhastCons 28way Placental Mammal with no overlap with RefSeq genes downloaded from UCSC on 15th June 2010 on HG18. Ranked according to LOD score to select the top-ranked, most-conserved elements

Downloaded in hg19 on 4 th August 2010 from UCSC. These were ranked acording to LOD score. Min LOD score was 397. Max LOD score was 3822.

**lincRNAs**
*Definition:* GENCODE annotation levels 1,2 and 3 on Hg19, downloaded on 22nd July 2010, including exons in transcripts annotated lincRNA

#### *Known typable untagged CNVs:*

**Rationale:** only target common CNVs that are not well-tagged in SNP chip data. Well-tagged CNVs can be tested for association indirectly using these tagging SNPs.

**Definition:**
1. clusterable by WTCCC+
2. classified as a 'good CNV' by WTCCC+ (which generally means high clustering quality and non-redundant)
3. Have $r^2$ of less than 0.5 with a SNP on the Illumina 660k chip

Originally on HG18.  Probes not lifted across to HG19 – original probe identifiers used.

#### *Control loci*

Agilent replicate probes (n=1000 probes replicated x5) include X probes (n=47 loci replicated x5) which will allow dose response to be measured on both slides.

**Sample tracking:**
30 common (MAF>0.4) CNVs that can be easily assayed (WTCCC posterior >0.98) on the Agilent aCGH platform with 10 probes per CNV and that are well-tagged  ($r^2$>0.99) by nearby SNPs on the Illumina 660k SNP chip that can also be assayed using Sequenom (28 of these assays are working in Sequenom).

Originally on HG18 – these have been lifted over to HG19.

***Table 1. Breakdown of targeted loci by type***

| Source | Number | Median size |
|---|---|---|
| Tagged CNVs (28 working on Sequenom) | 30 | 1370 |
| Untagged CNVs | 1,662 | 4492 HG18 |
| GENCODE protein-coding transcripts + miRNAs (merged) | 224,508 | 134 |
| Enhancers | 622 | 1580 |
| Highly-conserved non-coding sequences | 19,378 | 370 |
| lincRNAs | 5,221 | 1163 |

#### Probe design:

##### Functional elements:

Attempt to design 5 unique probes per functional element with probe score >0.4. Consider the sequence to be targeted to be an interval defined by the functional element ±200bp on either side

If more than 5 probes available prioritise probes on the basis of Agilent *in silico* probe performance score, while minimizing overlap between probes.

If fewer than 5 probes can be found replicate the highest scoring probe(s) until 5 probes

Probes selected from a set of probes drawn from (i) eARRAY catalog downloaded on 5 th July 2010 and (ii) tiling probes across the target region (element ±200bp) on both strands

The probe set picked using these parameters was then edited.

To distribute probes more evenly within targeted regions, regions were more than half the region was empty were found. Probes within these unbalanced regions were given a score based on how well they even out the coverage of the region.  These new probes were added to the regions. In some cases lowscoring probes were replaced to make room for the new probe. Probes were added without replacement where the Agilent quality score of the new probe was more than 0.3 below the worst probe, or where removal of a probe would itself cause a similar imbalance.

When a probe is picked the overlap threshold is reset to zero. Then it is incremented by one until a probe is picked. This means that the probe picked will be the first to pass the overlap threshold, not necessarily the highest scoring probe, although it will be the highest scoring of those that pass the threshold.

To increase the scores of probes in targeted regions extra copies were transferred to a higher scoring overlapping probe.

**Known typable untagged CNVs:**
IDs of probes targeting these CNVs selected from the WTCCC+ array design and mapped onto HG19 using BLAT on the probe sequence.

The untagged cnv probe set has 15428 probes. There are 14745 unique probes. Of these 8006 "map" to hg19. Unmapped probes are present on the array on the same slide as the mapped probes for that CNV.

(Mapping:- blat output is converted to ucsc style output (with span, identity and score), the best scoring hit is chosen provided:

Score == qSize (so that the size of the hit is the size of the probe)

Identity == 100 (so that there is a perfect match)

The chr matches the consensus chr for that CNV

The consensus chr for each cnv was derived by taking the best hit for each probe and a vote for each CNV. In some cases the there was a 50:50 split most likely chr was selected by hand using second best hits.

Probes in untagged CNV’s were also assiged to chromosomes using the Agilent hg19 probe location, if this could be obtained from the eARRAY catalog downloaded on 5 th July 2010. Locations were assigned using the Agilent location. If the Agilent mapping was unavalible or inconsistent then the BLAT mapping was used. If both these methods failed then the probes in that CNV were assigned to chr0 and placed on array1.

**Backbone probes:**
Probes selected from eARRAY catalog downloaded on 5 th July 2010. Selected to generate regular coverage while not selecting probes with low design scores.

The gaps in coverage were calculated genome-wide. The algorithm aims to place a feature in the centre of the largest gaps. Each time a feature is placed the gap is split by this feature into two new gaps. This algorithm works on the 100 largest gaps in parallel to improve performance and then selects the next 100 largest from both the un-touched gaps and pairs of new gaps just created.

To bias the selection towards the centre of the gap, each feature within the gap is assigned a gap score composed of the score of the feature multiplied by a measure of its centrality. The maximum of the proportional division of the gap by the feature is squared and used as the centrality measure.

**Control loci (patient tagging assays):**
The ten top scoring probes for each control CNV region were selected. (HG19) There are 10 probes per CNV.

***Table 2. Breakdown of probes designed by type of target***

| Target type | # targets | # targets with 5 unique probes (%) | # targets with 5 probes | # targets with 0 probes (%) | # probes (%) | Median probe score |
|---|---|---|---|---|---|---|
| GENCODE protein-coding transcripts + miRNAs (merged) | 224,508 | 207,980 (92.638%) | 216,584 (96.471%) | 6,801 (3.029%) | 1,108,588 (58.295%) | 0.7648 |
| Highly-conserved non-coding sequences | 19,378 | 18751 (96.764%) | 19098 (98.555%) | 23 (0.119%) | 97,786 (5.142%) | 0.9277 |
| lincRNAs | 5,221 | 4809 (92.109%) | 5114 (97.951%) | 60 (1.149%) | 37,098 (1.951%) | 0.875 |
| Enhancers | 622 | 620 (99.678%) | 620 (99.678%) | 1 (0.161%) | 3,265 (0.172%) | 0.9686 |
| Untagged CNVs | 1,662 | *NA* | | *NA* | 15,428 (0.811%) | 0.92550 |
| Tagged CNVs | 30 | *NA* | | *NA* | 300 (0.016%) | 0.9634 |
| Backbone | *NA* | *NA* | | *NA* | 665,801 (35.011%) | 0.9735 |
| **TOTAL** | 249,759 | 232,190 *(92.966%)* | 241,446 (96.672%) | 6,885 (2.757%) | 1,901,686 | 0.93110 |

**Apportioning of chromosomes per slide:**

*Array 1*
*Chromosomes: 1, 3, 5, 7, 9, 12, 15, 21, 22, X, Y*

*Array 2*
*Chromosomes: 2, 4, 6, 8, 10, 11, 13, 14, 16, 18, 19, 20*

***Table 3. Breakdown of probes on each array by chromosome***

| Chromosome | # probes slide 1 | # probes slide 2 |
|---|---|---|
| **chr1** | 172389 | 1382 |
| **chr2** | 1471 | 157069 |
| **chr3** | 125763 | 1201 |
| **chr4** | 1164 | 99169 |
| **chr5** | 102366 | 1061 |
| **chr6** | 1151 | 104490 |
| **chr7** | 98172 | 1028 |
| **chr8** | 952 | 80429 |
| **chr9** | 77314 | 726 |
| **chr10** | 838 | 82260 |
| **chr11** | 897 | 97798 |
| **chr12** | 97945 | 791 |
| **chr13** | 647 | 49324 |
| **chr14** | 549 | 60772 |
| **chr15** | 59969 | 564 |
| **chr16** | 848 | 65990 |
| **chr17** | 82537 | 507 |
| **chr18** | 514 | 40530 |
| **chr19** | 358 | 77124 |
| **chr20** | 386 | 44365 |
| **chr21** | 20887 | 252 |
| **chr22** | 31937 | 215 |
| **chrX** | 80580 | 235 |
| **chrY** | 6152 | 5 |
| **unmapped** | 1545 | 44 |
| **total** | 967331 | 967331 |

**Note:** Agilent normalization probes (on all autosomes) and well-tagged CNVs for sample tracking are present on both arrays, hence not all probes on a given chromosome are specific to one slide.
